# The Molecular Human – A Roadmap of Molecular Interactions Linking Multiomics Networks with Disease Endpoints

**DOI:** 10.1101/2022.10.31.22281758

**Authors:** Anna Halama, Shaza Zaghlool, Gaurav Thareja, Sara Kader, Wadha Al Muftha, Marjonneke Mook-Kanamori, Hina Sarwath, Yasmin Ali Mohamoud, Sabine Ameling, Maja Pucic Baković, Jan Krumsiek, Cornelia Prehn, Jerzy Adamski, Nele Friedrich, Uwe Völker, Manfred Wuhrer, Gordan Lauc, Hani Najafi, Joel A Malek, Johannes Graumann, Dennis Mook-Kanamori, Frank Schmidt, Karsten Suhre

## Abstract

In-depth multiomics phenotyping can provide a molecular understanding of complex physiological processes and their pathologies. Here, we report on the application of 18 diverse deep molecular phenotyping (omics-) technologies to urine, blood, and saliva samples from 391 participants of the multiethnic diabetes study QMDiab. We integrated quantitative readouts of 6,304 molecular traits with 1,221,345 genetic variants, methylation at 470,837 DNA CpG sites, and gene expression of 57,000 transcripts using between-platform mutual best correlations, within-platform partial correlations, and genome-, epigenome-, transcriptome-, and phenome-wide associations. The achieved molecular network covers over 34,000 statistically significant trait-trait links and illustrates “The Molecular Human”. We describe the variances explained by each omics layer in the phenotypes age, sex, BMI, and diabetes state, platform complementarity, and the inherent correlation structures of multiomics. Finally, we discuss biological aspects of the networks relevant to the molecular basis of complex disorders. We developed a web-based interface to “The Molecular Human”, which is freely accessible at http://comics.metabolomix.com and allows dynamic interaction with the data.

## INTRODUCTION

The quote *“Learn how to see. Realize that everything connects to everything else”* by Leonardo Da Vinci becomes substantive in the context of high-throughput deep molecular phenotyping technologies that enable the measurement of hundreds or even thousands of quantitative readouts of the genome, transcriptome, proteome, metabolome, and glycome as well as related intermediate omics layers, such as the epigenome, and microRNA-ome (Chen et al., 2012; Dai et al., 2021; Hasin et al., 2017; Karczewski and Snyder, 2018; Sailani et al., 2020; Suhre et al., 2010)(de Haan et al., 2022). Integrated into a single study, these readouts simultaneously provide insight into the molecular interactions that define the physiological and the pathophysiological processes in the human body.

Deep molecular phenotyping at large-scale using multiple platforms and matrices (“multiomics”) in large cohort studies is becoming increasingly attractive and is already being driven by the UK Biobank consortia, which genotyped 500,000 participants and are currently acquiring transcriptomics, proteomics, and metabolomics data for a large fraction of them. With many different technologies and platforms available, however, questions arise as to the choice of platform, platform complementarity, and most importantly, how to integrate the disparate and complex data sets once they have been captured as well as how to visualize and present the results outcomes.

Here, we report on what is arguably one of the most deeply phenotyped cohort studies to date. The Qatar Metabolomics Study of Diabetes (QMDiab) (Mook-Kanamori et al., 2014) was originally designed as a diabetes case-control study in the multiethnic population of Qatar. We collected multiple aliquots of blood, urine, and saliva samples from 391 volunteers, with and without diabetes, of predominantly Arab, Filipino, or Indian ethnic backgrounds with the goal of acquiring sufficient material for multiomic analysis (see methods). The collected samples were subsequently characterized on 18 different high-thruput omics platforms. These included blood circulating micro-RNAs, proteins, molecular levels of IgG- and IgA-glycosylations, N-glycosylation of total protein, metabolites in urine, saliva and plasma measured on targeted and non-targeted Nuclear Magnetic Resonance (NMR)- and mass spectrometry (MS)-based metabolomics platforms, and lipid composition by size-resolved lipo-proteomics as well as complex lipid profiles. Over 6,300 individual omics data points were collected for each study participant. In addition, samples were genotyped for 1.2 million genetic variants, the white blood cell transcriptome was sequenced at a depth of 20 Mio reads to quantify the expression of 57,000 transcripts, and DNA methylation levels for 450,000 CpG sites were determined.

We aimed to simultaneously answer technical questions related to omics platform complementarity, data integration, visualization, and accessibility, as well as biological questions related to interrelationships between these molecular traits and their association with complex disease. The ultimate goal was to draw, through these omics’ layers, an image of “The Molecular Human”.

To achieve this goal, we connected all multiomics traits using appropriate measures, that is, mutual best hits (MBH) of between-platform correlations, partial correlations to construct Gaussian Graphical Models (GGMs) within individual omics-layers, and genome-wide (GWAS), epigenome-wide (EWAS) and transcriptome-wide (TWAS) associations between the high-dimensional genomics readouts and the other omics layers. Finally, we integrated all connections into a multiomics network with clinical endpoints through phenome- and genome-wide disease associations. We evaluate each omics layer for its potential to explain the variability of the study participants’ age, sex, BMI and diabetes state, and further quantified the between layer degree of shared mutual information. Finally, we present three distinct use cases to show the generalizability of the approach. To facilitate rapid sharing of our results, and also to provide the user with the possibility of testing the interactions of their own molecules of interest in the context of other omics layers, we developed a webserver called *Connecting Omics* (COmics) (http://comics.metabolomix.com). We also provide the full network in digital format (Cytoscape) for free download.

## RESULTS

### Deep molecular phenotyping of 391 individuals using 18 omics platforms in three sample matrices

Urine, saliva, and blood samples from 391 subjects in the QMDiab study were analyzed on 18 technically distinct platforms (see **Table 1** for platform abbreviation) relying on sequencing-, microarray-, MS-, NMR-, affinity binding-, chromatography-, and biochemistry assay-based technologies (see Methods, **Table 1, Figure 1, and Supplementary Table 1** for all molecules measured on non-genomics platforms). The number of quantitative molecular traits determined by the non-genomics platforms ranged from 36 to 1,201, and the number of samples shared between two platforms from 229 to 356 **(Table 2)**. In total, we determined quantitative measures for up to 6,304 molecular traits per sample along with genotypes for 1,221,345 autosomal SNPs, expression levels of 57,773 transcripts, and DNA methylation of 470,837 CpG sites.

**Table 1.** Overview on applied omics technologies.

| Omics | Measurement | Technique/Platform | Matrix | Label |
| --- | --- | --- | --- | --- |
| **GENOMICS** | Genotype | Infinium Human Omni 2.5-8 v1.2 BeadChip kit | DNA extracted from buffy coat fraction from whole blood | DNA |
| **METHYLOMICS** | DNA methylation | Illumina Infinium HumanMethylation450 BeadChip kit | DNA, same as for genomics | MET |
| **TRANSCRIPTOMICS** | Gene expression | RNA-sequencing based Illumina ~20M reads | RNA extracted from PaxTube | RNA |
|  | microRNA expression | microRNA profiling based multiplex qPCR, Exiqon | RNA extracted from EDTA plasma | miRNA |
| **PROTEOMICS** | Protein abundance | Slow Off-rate Modified Aptamer (SOMAmer), Somalogic 1,1k | EDTA plasma | SOMA |
|  | Relative protein abundance | Proximity Extension Assay (PEA) based Olink Target 96 Metabolism & Cardiometabolism panels | Heparin plasma | OLINK |
| **GLYCOMICS** | Total plasma N-glycosylation | Hydrophilic interaction ultra-performance liquid chromatography | EDTA plasma | PGP |
|  |  | (HILIC-UPLC) based Genos pipeline |  |  |
|  | IgG glycosylation | Liquid chromatography mass spectrometry (LC-MS) based Genos pipeline | EDTA plasma | IgG |
|  | IgA & IgG glycosylation | LC-MS based (Momcilovic et al., 2020) pipeline | EDTA plasma | IgA |
| **LIPOPROTEOMICS** | Lipoproteins | Proton nuclear magnetic resonance (<sup>1</sup>H NMR) based Nightingale technology | EDTA plasma | BRAIN |
| **LIPIDOMICS** | Lipid concentration | LC-MS based on Lipidzyzer technology at Metabolon | EDTA plasma | LD |
|  | Lipids and other metabolite concentration | Flow injection analysis (FIA)- MS based Biocrates technology | EDTA plasma | BM |
| **METABOLOMICS** | Metabolite level | (HILIC-MS) & (UPLC-MS) based HD4 Metabolon | EDTA plasma | HDF |
|  | Metabolite level | Gas chromatography (GC)-MS (UPLC-MS) based HD2 Metabolon | EDTA plasma | PM |
|  | Metabolite level | (GC-MS) & (UPLC-MS) based HD2 Metabolon | Saliva | SM |
|  | Metabolite level | GC-MS & UPLC-MS based HD2 Metabolon | Urine | UM |
|  | Metabolite level | <sup>1</sup>H NMR deploying Chenomx for annotation, based on (Budde et al., 2016) pipeline | Urine | CM |
| **CLINICAL** | Clinical biochemistry and blood counts | Cobas 6000; Roche Diagnostics | Blood/Urine | CLIN |

**Table 2:** Mutual best hits (MBH) between platforms. The upper triangle of this matrix indicates the number of mutual best hits identified between the respective platforms, the diagonal contains the number of traits evaluated for that platform, and the lower triangle reports the number of samples for which data was available for both platforms in parallel. Platform abbreviations are explained in Table 1.

|  | miRNA | SOMA | OLINK | PGP | IgG | IgA&IgG | BRAIN | UM | PM | SM | HDF | BM | CM | LD | RNA | CLIN |
| --- | --- | --- | --- | --- | --- | --- | --- | --- | --- | --- | --- | --- | --- | --- | --- | --- |
| miRNA | 169 | 6 | 2 | 1 | 0 | 1 | 2 | 0 | 2 | 0 | 7 | 0 | 2 | 0 | 0 | 2 |
| SOMA | 337 | 1129 | 73 | 14 | 8 | 19 | 20 | 26 | 32 | 0 | 36 | 17 | 5 | 18 | 12 | 16 |
| OLINK | 309 | 323 | 184 | 10 | 1 | 7 | 18 | 12 | 18 | 0 | 23 | 20 | 1 | 15 | 8 | 12 |
| PGP | 326 | 344 | 313 | 36 | 10 | 14 | 7 | 3 | 8 | 0 | 7 | 4 | 2 | 4 | 4 | 6 |
| IgG | 326 | 340 | 310 | 331 | 60 | 31 | 5 | 4 | 4 | 0 | 7 | 2 | 2 | 3 | 3 | 4 |
| IgA&IgG | 325 | 341 | 322 | 330 | 291 | 178 | 5 | 9 | 9 | 2 | 11 | 7 | 3 | 4 | 2 | 7 |
| BRAIN | 333 | 350 | 317 | 339 | 337 | 335 | 224 | 15 | 19 | 1 | 28 | 18 | 5 | 8 | 1 | 10 |
| UM | 314 | 331 | 301 | 319 | 316 | 319 | 325 | 805 | 174 | 14 | 214 | 26 | 43 | 22 | 2 | 8 |
| PM | 321 | 339 | 308 | 327 | 323 | 326 | 333 | 347 | 600 | 24 | 369 | 45 | 21 | 43 | 3 | 15 |
| SM | 254 | 267 | 242 | 258 | 250 | 252 | 262 | 273 | 281 | 434 | 21 | 1 | 7 | 0 | 0 | 4 |
| HDF | 295 | 308 | 290 | 300 | 299 | 306 | 307 | 285 | 292 | 229 | 1020 | 75 | 21 | 96 | 1 | 14 |
| BM | 319 | 337 | 306 | 326 | 322 | 324 | 331 | 345 | 356 | 279 | 290 | 163 | 5 | 44 | 0 | 6 |
| CM | 308 | 324 | 296 | 313 | 310 | 313 | 318 | 353 | 340 | 269 | 281 | 338 | 60 | 4 | 1 | 4 |
| LD | 310 | 323 | 303 | 313 | 312 | 320 | 322 | 300 | 307 | 241 | 302 | 305 | 293 | 1201 | 0 | 5 |
| RNA | 296 | 311 | 287 | 301 | 299 | 301 | 306 | 297 | 300 | 235 | 270 | 299 | 291 | 285 | 1239* | 7 |
| CLIN | 321 | 339 | 307 | 327 | 323 | 323 | 332 | 358 | 357 | 274 | 293 | 355 | 351 | 306 | 304 | 41 |

\*Note: Genotype (DNA) and methylation (MET) data were not included in the MBH computation. Transcriptome (RNA) was limited to 1,239 transcripts that are also covered by the two proteomics platforms (SOMA, OLINK).

**Figure 1.**
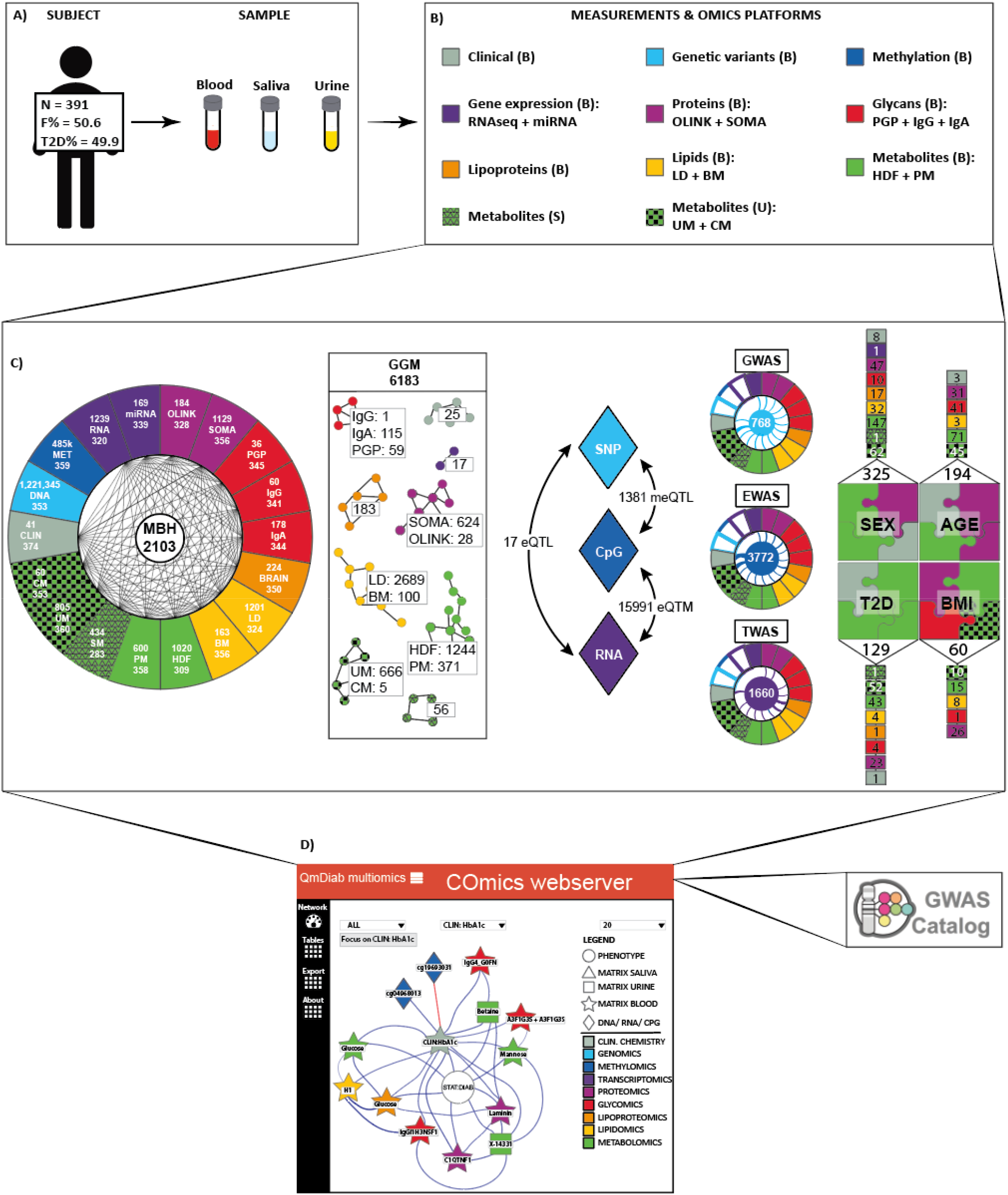
Overview on the subject and data sets. **A)** Collected data and omics platforms used for data generation; **B)** Mutual best hit describing the significant associations identified per each omics platform. **C)** Multiomics GWAS, multiomics EWAS, and multiomics TWAS; **D)** Molecular network of interactions constructed after computation of within-trait partial correlations available in the form of freely accessible server (https://littleswissriver.shinyapps.io/generate_network/) linking molecular networks with the genetic disease risks via GWAS server). CLIN: clinical chemistry parameters; DNA: genotype data of ; MET: DNA methylation sites; RNA: RNA transcripts determined with RNA-sequencing; miRNA: microRNA profiles; SOMA: blood circulating proteins measured with aptamer-based technology (SomaLogic); OLINK: blood circulating proteins measured using high-multiplex immunoassays (Olink); PGP: glycan traits N-glycosylation; IgG: IgG-glycopepdides; IgA: IgA and IgG-glycopeptides BRAIN: plasma lipoproteins; LD: plasma lipids quantified using Lipidyzer; BM: plasma lipids quantified with Biocrates p150 kit; HDF: plasma metabolic traits profiled on HD4 platform (Metabolon); PM: plasma metabolic traits profiled on HD2 platform (Metabolon); SM: saliva metabolic traits profiled on HD2 platform (Metabolon); UM: urine metabolic traits profiled on HD2 platform (Metabolon); CM urine metabolites quantified with ^1^H NMR deploying Chenomx.

Complex correlation structures within and between platforms pose major challenges to the integration of these datasets. For example, correlation between complex lipid species may be driven by the abundance of common precursor fatty acids, but also by factors determining interconversion between different classes. To cope with these challenges, and based on prior experience (Krumsiek et al., 2011; Suhre et al., 2017), we adopted a strategy using MBHs correlation between platforms, partial correlations within platforms, and linear model associations for genomics traits (GWAS, EWAS, and TWAS hits). In total we identified 2,103 unique MBHs, 6,183 partial correlations, as well as 768 GWAS, 3,772 EWAS, and 1,660 TWAS hits at stringent Bonferroni significance levels (see methods).

To simplify data access and result visualization we integrated all associations along with GWAS catalogue information into a molecular network and constructed a molecular network consisting of in excess of 34,000 edges and 6,304 nodes, which we made available in the form of the freely accessible *COmics* webserver (http://comics.metabolomix.com). All associations and the server software are available for download as source code and a Docker image (https://github.com/karstensuhre/comics).

### Information content of the platforms: explained variance of age, sex, BMI, and diabetes state

The molecular composition of the body at different omics layers is usable to explore effects of sex (Krumsiek et al., 2015; Miike et al., 2010; Singmann et al., 2015), measure biological age (Kristic et al., 2014)(Bocklandt et al., 2011; Hertel et al., 2016; Lehallier et al., 2019; Peters et al., 2015; Robinson et al., 2020; Tanaka et al., 2018), or study diabetes progression (Pena et al., 2016; Schrader et al., 2022; Wang-Sattler et al., 2012). Here we investigated which of the molecular traits and platforms most accurately characterize phenotypes such as age, sex, BMI and type 2 diabetes (T2D). First, we determined molecules associated with the phenotypes age, sex, BMI and T2D and identified 194, 325, 60, and 129 associated molecules, respectively **(Supplementary Table 2 - 5)**. Next, we examined the percentage of age, sex, BMI and T2D variance, which may be explained in data from each individual platform. We trained a random forest model for two continuous (age and BMI) and two dichotomous traits (sex and diabetes state) on each platform and estimated the variation explained by the respective omics phenotype **(Table 3)**. We found that both metabolomics and proteomics accurately describe the variation of all the investigated phenotypic traits. For instance, the variations in sex (95%) and T2D (86%) were most precisely captured by the HDF platform, age (54% and 52%) by the OLINK and SOMA platforms respectively, and BMI (42%) by the SOMA platform. The molecules measured on clinical chemistry data (CLIN) were accurate towards age (55%), sex (93%) and T2D (92%).

**Table 3.** Percentage of the variance explained in age, sex, BMI and diabetes state by platform.

|  | AGE [%] | SEX [%] | BMI [%] | DIAB [%] |
| --- | --- | --- | --- | --- |
| CLIN | 54.9 | 92.5 | 17.7 | 92 |
| RNA | 24.6 | 70.6 | 9.4 | 67.2 |
| miRNA | 9.7 | 61.4 | 3.2 | 58.4 |
| OLINK | 53.8 | 85.4 | 22.1 | 79.6 |
| SOMA | 52.4 | 93.3 | 41.7 | 82.3 |
| PGP | 44.2 | 73.3 | 26.6 | 72.5 |
| IgG | 46.3 | 63.9 | 4.3 | 71.8 |
| IgA | 45.1 | 73 | 7 | 73 |
| BRAIN | 28.7 | 77.4 | 16.5 | 76.6 |
| LD | 26.4 | 76.8 | 18.9 | 68.2 |
| BM | 22.4 | 77.5 | 23.9 | 83.1 |
| HDF | 50.9 | 95.1 | 28.6 | 86.4 |
| PM | 51.7 | 91.6 | 24.7 | 86.3 |
| SM | 20.7 | 64.3 | 5.1 | 71 |
| UM | 50.3 | 89.4 | 27.3 | 81.9 |
| CM | 37 | 83.8 | 16.3 | 81.9 |

This data identifies individual platform capability to explain variance in age, sex, BMI and T2D phenotypes

### Mutual Best Hits between platforms assessment

MBHs, also known as reciprocal or bidirectional best hits, are a hypothesis-free approach to identify molecular orthologs between platforms. Inspired by the use of MBHs to distinguish orthologous from paralogous genes in bacterial genome analysis (Overbeek et al., 1999; Tatusov et al., 2001), we use this approach here to identify ortholog readouts between two platforms. This can be challenging when the platforms capture related features using different techniques and at varying depth. Examples are differences in resolution of lipid side chains or protein glycosylation. Examining these individual MBHs pairs may also provide insight into the measurement quality of the specific molecule on each individual platform, reveal potential issues with molecule annotations, and help define the general overlap between platforms.

The number of MBHs between every two platforms (⇔) is presented in **Table 2**, and the correlation levels of all statistically significant MBHs are provided in **Supplementary Table 6**. The number of traits determined by each platform varies, so the relative information content provided by one platform compared to another is also different. For example, 60 urine metabolites were measured using the NMR-based Chenomx platform (CM; see **Table 1 for platform abbreviations**) and 805 molecules were quantified on the MS-based Metabolon HD2 platform (UM). We identified 43 significant MBHs between the two platforms, accounting for 72% of the traits determined by the CM platform but only 5% by the UM platform.

Given the technological similarity, the largest number of MBHs was found between successive generations of the Metabolon platforms (HDF ⇔ PM) with 369 hits. Out of these, 291 paired identically annotated metabolites, 57 MBHs linked an unknown metabolite measured on the older platform to an annotated molecule measured on the more recent platform (e.g. X-18601 ⇔ androstenediol (3beta,17beta)-monosulfate), and 21 MBHs linked apparently differing molecules in related pathways (7 molecules; e.g. threonate ⇔ oxalate) or unknowns (14 molecules). As the Metabolon platforms differ with respect to the technology employed, this shows a robust concordance of platform performance and progressing component identification over time.

Detailed IgG glycosylation was determined by two independent platforms depicted as IgG and IgA. In the results, 29 out of 31 identified MBHs mapped to the same glycan structure, showing excellent agreement between both platforms.

Two different affinity proteomics platforms were used, one based on aptamers (SOMA, 1129 traits), the other based on antibody pairs as binders (OLINK, 184 traits). Out of 72 proteins that overlapped between both platforms, 52 were linked by MBH. We further investigated the correlations of overlapping proteins **(Supplementary Table 7)**. We further focused on those proteins, which were not captured by a MBH and showed low correlation to further explore its impact on molecular network **(Supplementary Information Note 1**). We found different molecular networks around those proteins which suggests that proteins showing low correlation should be validated with alternative technical platform to ensure the correctness of the measurement.

Overall, the technologically different platforms, deployed to cover same omics, display concordance in respect of detected molecules, which underscores good quality of selected methods.

### Evaluation of platform performance through the strength of GWAS hits

The strength of a genetic association depends on sample number, the effect size, as well as the technical and biological variability of the phenotype. Replication of genetic signals across platforms provides an independent assessment of the strength of that platform, especially when evaluated on sample aliquots from the same study, where technical variability is the only factor that differs between platforms. Thus, comparing association p-values for QTLs with different omics phenotypes on an identical genetic variant provides an objective measure for comparing readouts from two platforms.

Exploiting this property **(see Supplementary Information Note 2)**, we found none of the platforms employed to generally outcompete the alternatives regarding strength of genetic association, but individual platforms showed superior performance for certain molecules.

### Deploying platform complementarity to provide further insight into the structure of complex lipids

The composition of fatty acid (FA) side chains in complex lipids such as phosphatidylcholine or triacyclglycerols play a role in a broad range of biological processes and it is thus critical to determine FA composition in measured lipids. This information is, however, not provided for phosphatidylcholines measured on the BM platform or triacyclglycerols measured on the LD platform. We previously showed that the composition of phosphatidylcholines measured on the BM can be resolved by LD platform (Quell et al., 2019). Here, we investigated whether MBH may support more accurate determination of the structural composition of complex lipids. Indeed, after examining MBH between the two lipidomics platforms (LD ⇔ BM) and between the metabolomics and lipidomics platforms (HDF ⇔ BM) and (HDF ⇔ LD), we resolved the FA side chain composition for a number of complex lipids. The side chain composition of phosphatidylcholines measured on the BM platform, for instance, were delineated using MBH (e.g. PC_aa_C32:1 ⇔ PC(16:0/16:1), PC_aa_C40:6 ⇔ PC(18:0/22:6)) **(Figure 2A)**, in line with our previous study (Quell et al., 2019). The characterization of fatty acid chains in triacylglycerol was similarly refined (TAG48:2-FA14:0 ⇔ myristoyl-linoleoyl-glycerol (14:0/18:2); TAG54:6-FA22:6 ⇔ palmitoyl-docosahexaenoyl-glycerl (16:0/22:6)) **(Figure 2B)**.

**Figure 2.**
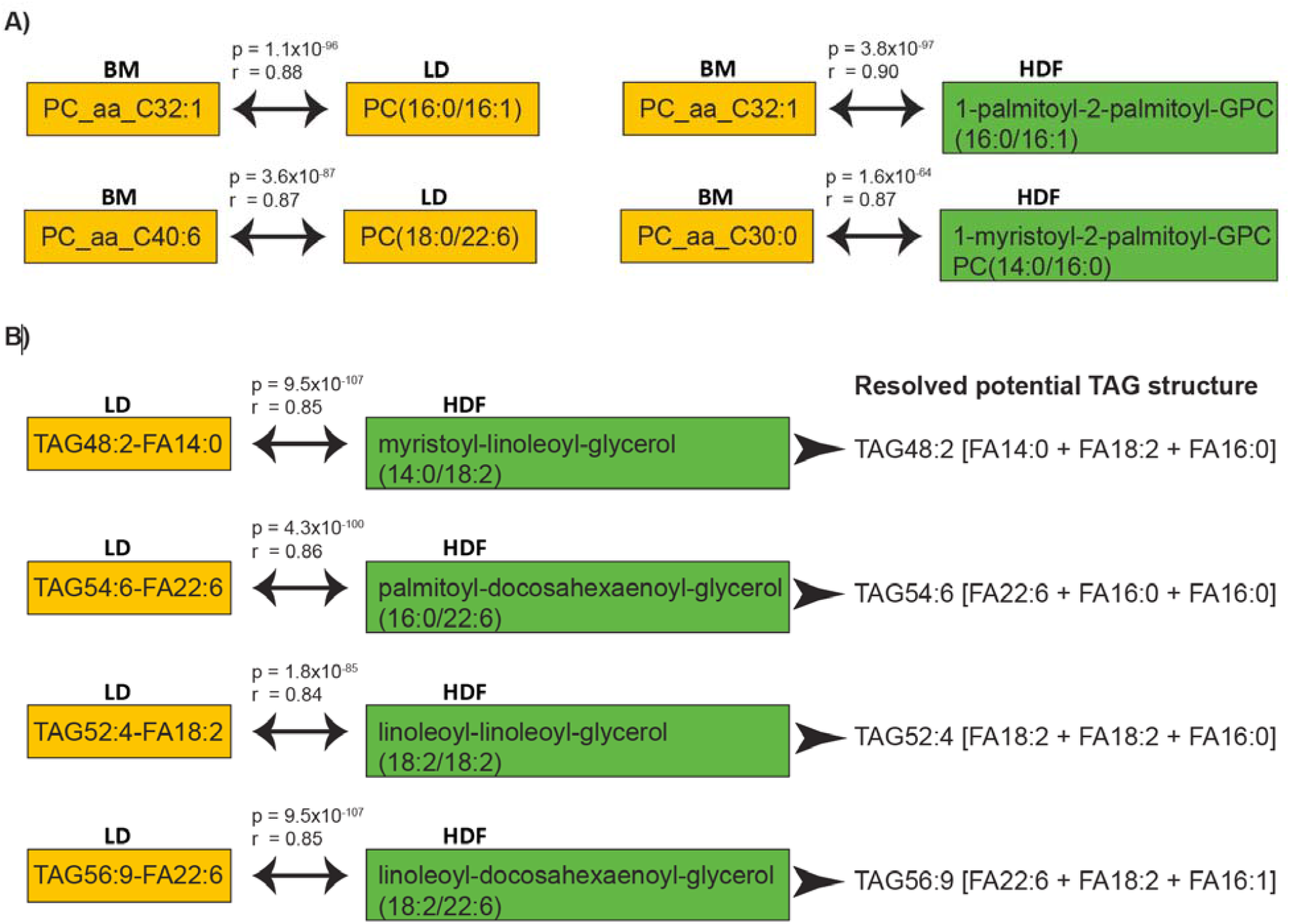
Omics platforms overlap and complementarity. **A) & B)** The structure of complex lipids was revealed with by complementary platforms connected with MBH.

These examples indicate how combining two technologically similar platforms can add valuable information, making them complimentary rather than redundant.

### Molecular associations within and between omics represent biological process

Connection within and between distinct omics layers may reveal biological interactions via the molecules showing association and various computational strategies may be applied to provide such insight. One such example are Gaussian Graphical Models (GGMs), based on partial correlation coefficients, which we previously employed to reconstruct pathways from metabolomics data (Krumsiek et al., 2011). GWAS with intermediate phenotypes such as metabolomics, proteomics, or epigenomics have already been numerously conducted across different populations providing insight into human physiology and various pathophysiological processes (Gilly et al., 2020; Huan et al., 2019; Kettunen et al., 2016; Suhre et al., 2021; Suhre et al., 2011). For instance, our previous EWAS lead to identification of associations between epigenetic variations and different biological traits (Li et al., 2019)(Zaghlool et al., 2020; Zaghlool et al., 2018).

Here we will discuss multiple examples showing how the computed associations between various omics, reflect on the underlying biology. This includes GGMs **(Supplementary Table 8);** MBH between omics (**Table 2** and **Supplementary Table 6**); associations of SNPs with methylation levels (meQTL’s) (1,381 meQTL’s see **Supplementary Table 9**); association of methylation levels with mRNA expression levels (eQTM’s) (15,991 eQTM’s see **Supplementary Table 10**); association of SNPs with mRNA expression (eQTL’s) (17 eQTL’s see **Supplementary Table 11**); GWAS with multiomics (mo) (768 omicQTLs at 586 independent genetic loci presented in **Supplementary Table 12**); EWAS with multiomics (moEWAS) (3,772 omicQTMs presented in **Supplementary Table 13**); and TWAS with multiomics (moTWAS) (1,660 omicQTRs presented in Supplementary Table 14). Prior to the evaluation of the biological relevance of computed associations, we checked whether the GWAS, EWAS, and TWAS resulted in the identification of previously reported hits. Our study replicates multiple previous findings as outlined in **Supplementary Information Note 3**. TWAS studies focus mainly on gene–trait associations from GWAS datasets (Gamazon et al., 2015; Gusev et al., 2016; Wainberg et al., 2019). To the best of our knowledge, this our moTWAS study is the first conducted so far. Examples highlighting biologically relevant findings are described below, and more general findings included in **Supplementary Information Note 3-7**.

### Examples of biological processes within various omics data captured by GGM’s

The number of identified partial correlations vary according to platform. For instance, 2,689 metabolite-metabolite associations were identified for the LD platform while only one glycan-glycan association was found for the IgG platform **(Supplementary Table 7)**. GGM associations help explain biochemical processes by connecting chemical reactions (e.g. association between cortisone ⇔ cortisol; fumarate ⇔ malate; glutamate ⇔ alpha-ketoglutarate). GGM also help in the understanding of physiological processes such as the association between Luteinizing hormone (LHB) and follicle stimulating hormone (FSHB), which synergistically stimulate follicular growth and ovulation (Filicori et al., 1999), as well as the metabolism and excretion of aspirin **(Figure 3A)**. Therefore, GGMs provide a simplified overview of the actual biological processes and can be graphically explored for each integrated omics layer in our COmics server.

**Figure 3.**
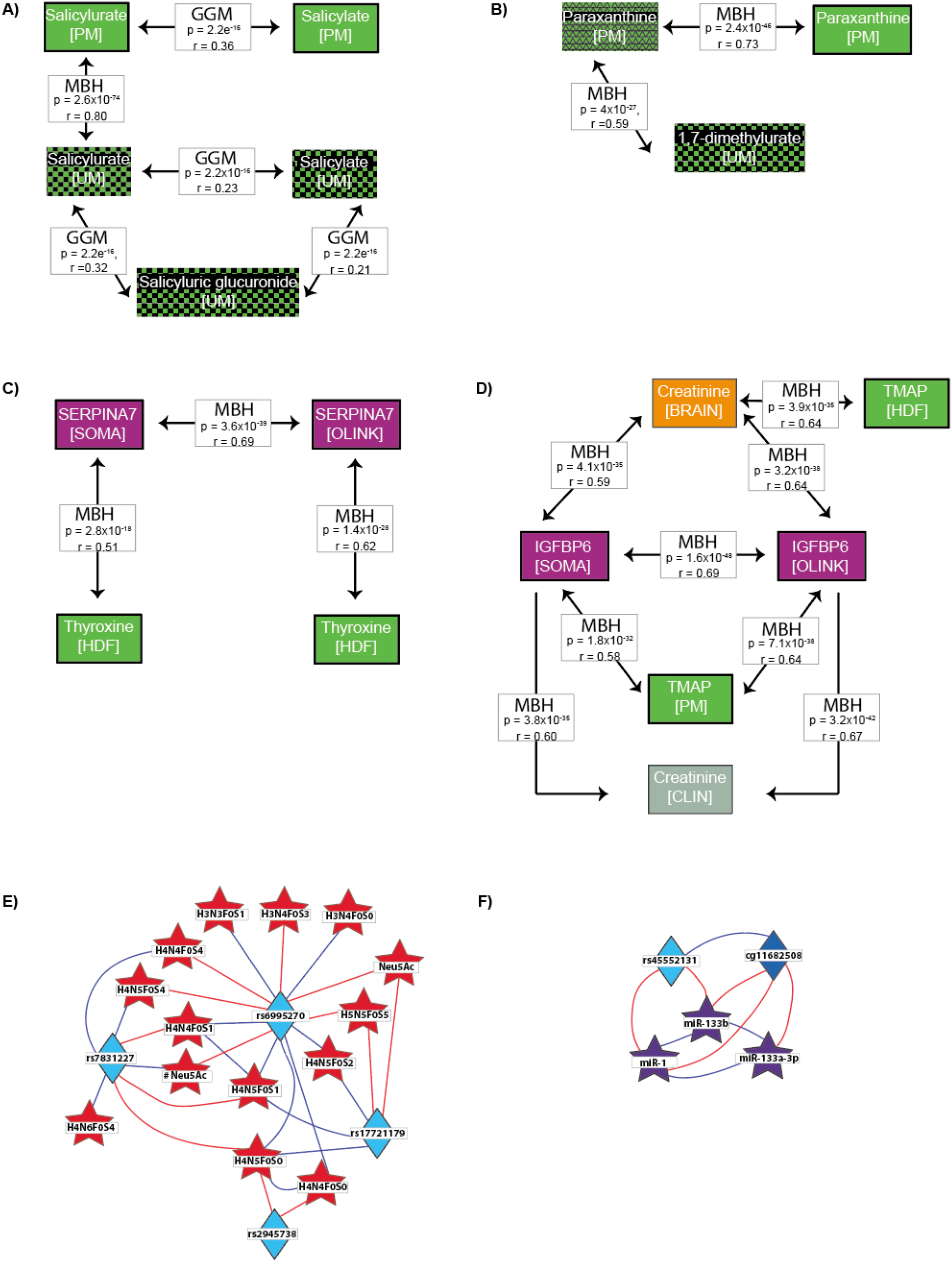
Biological processes reflected by connected omics with various strategies including: **A)** Gaussian Graphical Model (GGM). Observed associations potentially reflect on aspirin metabolism and excretion where salicylate is conjugated in the liver to form salicylurate and further metabolized to salicyluric glucuronide (Needs and Brooks, 1985); **B)** Between various matrix (saliva, blood, urine) Mutual Best Hit (MBH); **C) & D)** Between omics MBH; **E)** glycome GWAS reveled association between ST3GAL1 variants and IgA1 glycosylation; **F)** miRNA GWAS.

### Example of crosstalk between metabolites of different matrices: urine, saliva and plasma

MBH between urine, saliva and plasma metabolites, all measured on the Metabolon HD2 platform, inform on crosstalk between these matrices. We found 174, 24, and 14 MBHs between urine and plasma metabolites, plasma and saliva metabolites, and urine and saliva metabolites, respectively. Most MBHs connected identical molecules, reflecting on homeostasis between saliva and plasma, as well as the detoxification processes that occur in the kidney. As for the MBHs we found between metabolites from different matrices, this can be used to inform on physiological metabolism as well as disease related pathological processes. For instance, caffeine metabolism may serve as an example showcasing metabolic interactions between saliva, blood, and urine **(Figure 3B)**. Paraxanthine is the main caffeine metabolite in the body (Lelo et al., 1989). We found a MBH between plasma and salivary paraxanthine, in addition to a MBH between paraxanthine in saliva and its metabolic product, 1,7-dimethylurate (Nehlig, 2018) in urine. This shows how data between different sample matrices can be integrated and interpreted.

### Examples of biological interactions identified by MBH

MBHs can represent simple relationships such as molecular transport as well as more complex relationships pertaining to disease. For example, we found a MBH between thyroxine and SERPINA7, a thyroxine-binding globulin, which in the bloodstream carries thyroxine and triiodothyronine into thyroid gland (Contreras et al., 1981). This MBH between protein and metabolite was found independently of proteomic platform used **Figure 3 C**. A further MBH association found between creatinine, a marker of kidney function, and Insulin-like growth factor-binding protein-6 (IGFBP6), previously identified at elevated levels in children with chronic renal failure (Powell et al., 1997) potentially sheds light on that pathophysiology. Interestingly, we also found a MBH between IGFBP6 and N,N,N-trimethyl-alanylproline betaine, which was recently shown to be a signature of compromised kidney function (Velenosi et al., 2019) **(Figure 3 D)**. For more examples of biological interactions captured by MBHs, see **Supplementary Note 4**. Associations represent the underlying biological processes emphasize the complementarity between platforms measuring different omics.

### Examples of biological interactions identified by novel GWAS association

A description of the GWAS association is included in **Supplementary Information Note 5 & 6**. While analyzing our GWAS hits, we found previously unreported GWAS associations between multiple variants near ST3GAL1 (rs6995270, rs17721179, rs2945738, rs13264936), sialic acid and 11 different glycans, which all include N-acetylneuraminic acid (sialic acid) **(Figure 3 E)**. ST3GAL1 is a glycosyltransferase that catalyzes the transfer of sialic acid from Cytidine-5⍰-monophospho-N-acetylneuraminic acid (CMP-sialic acid) to galactosyl β(1,3)-N-acetylgalactosamine] Galβ1-3GalNAc (Lin et al., 2021; Wu et al., 2018). Previous studies suggested that ST3GAL1-mediated sialylation plays a role in the cancer cell strategy to evade immune attack making ST3GAL1 a potential treatment target (Lin et al., 2021), on which our study expands. With this example we further underscore importance of molecular traits like e.g. glycans, which serve as intermediate phenotypes for various pathological conditions and thus provide insight on genetic background and potential mechanistic link to clinical outcomes.

### Examples of biological interactions identified by GWAS and EWAS with microRNA

Genetic and epigenetic association with miRNA may provide further insight into regulatory mechanisms of microRNA transcription and elucidate roles of microRNA in mediating complex disease (Huan et al., 2015). We identified 10 mirQTLs of which four (miR-181a-5p, miR-133a-3p, miR-133b, and miR-1) replicated previous findings (Huan et al., 2015; Nikpay et al., 2019). Interestingly, we also observed association between three miR’s (miR-133b,miR-1, and miR-133a-3p), among which two (miR-133b and miR-1) showed association with rs45552131 (near C20orf166-AS1), and all three with cg11682508 (C20orf166) **(Figure 3F)** potentially suggesting their interplay in a regulatory process. The observed association between cg11682508 and rs45552131 replicates previous findings (Bonder et al., 2017). Expression of miR-1 and miR-133a is modulated by insulin and may be involved in insulin signaling. Given that both miRNA’s are derived from introns of protein-coding transcripts (C20orf166) (Granjon et al., 2009) it may be reasoned that cg11682508 identified here is also involved in insulin signaling. Interestingly, cg11682508 was previously described as one of the methylation sites being dysregulated in pancreatic islets of T2D subjects (Volkmar et al., 2012). Additional interesting findings from our EWAS can be found in **Supplementary Information Note 7**. This example shows not only replication of previous findings but is pointing towards novel CPG – miRNA axis.

### Examples of biological interactions identified by TWAS associations

The vast majority of identified TWAS associations (1,114 out of 1,660) were found with lipids and lipoproteins, while only a few with proteins (300) and metabolites (157). Of these the association between ENSG00000254415_SIGLEC14 ⇔ SIGLEC14 (SOMA), and ENSG00000115523_GNLY ⇔ GNLY (SOMA), for example, reflect on translation processes, where the expressed gene serves as a guide for the protein synthesis. Other associations like the one found between prednisolone, a corticosteroid, and the FKBP5 gene transcript previously described as elevated under oral corticosteroid (Bigler et al., 2017), may indicate medication-triggered treatment responses.

Our lipidomics TWAS (lTWAS) results feature prominently a group of five molecules with extensive networks of associations: GATA2 with 590 lipids, HDC with 516 lipids, FCER1A with 36 lipids, PDK4 with 18 lipids, as well as MS4A3 with 18 lipids. An overlap between molecules associated with gene transcripts of GATA2, HDC, MS4A3, and FCER1A but not with PDK4 **(Figure 4A)** reproduces ingenuity pathway analysis (IPA) that suggests potential interaction between GATA2, HDC, MS4A3, and FCER1A but not PDK4 **(Figure 4B)**. Lipids associated with PDK4 were largely fatty acids with various chain lengths **(Figure 4C)**. Changes in PDK4 expression were shown to play a role in lipid-related metabolic adaptation by stimulating fatty acids oxidation (Pettersen et al., 2019), which may explain the observed associations. GATA2, HDC, MS4A3 and FCER1A as group showed positive association with various high-density (HDL) and low-density lipoproteins (LDL), as well as negative association with triacylglycerols (TAG) and very-low-density lipoproteins (VLDL). GATA2 and HDC additionally showed positive association with phosphatidylcholines, lysophosphatidylcholines and apolipoprotein A (APOA) as well as negative association with diacylglycerols and apolipoprotein E (APOE) **(Figure 4D)**. The negative association between both GATA2 and HDC and TG as along with the positive association between GATA2 and HDC and HDL have been reported previously (Inouye et al., 2010). They may indicate a role of GATA2 and HDC gene transcripts in regulation of APOA and APOE metabolism, as well as a potential involvement in the pathology of various diseases related to dysregulated lipid metabolism such as e.g. cardiovascular diseases. Please see **Supplementary Information Note 8** for additional examples describing our EWAS findings.

**Figure 4.**
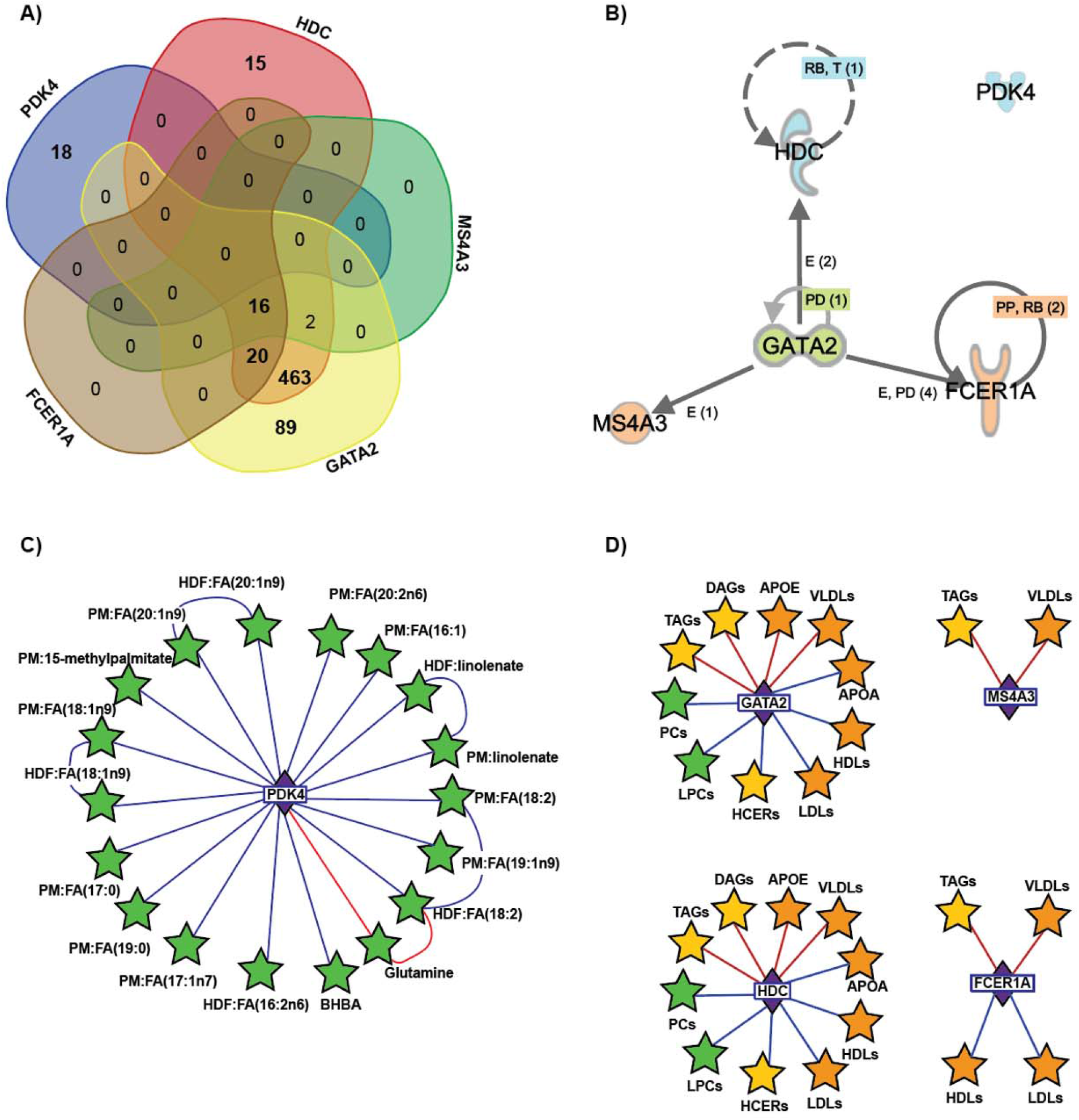
TWAS reveal interplay between GATA2, HDC and FCER1A but not PDK4. **A)** Venn diagram showing an overlap between molecules associated with gene transcripts of GATA2, HDC, MS4A3, and FCER1A but not PDK4. **B)** Ingenuity pathway analysis (IPA) revealed potential interaction between GATA2, HDC, MS4A3, and FCER1A but not PDK4. **C)** The molecules associated with PDK4. **D)** Associations between lipids structures and GATA2, HDC, MS4A3 and FCER1A.

### The Molecular Human - insight into architecture of complex diseases via COmics server

Diseases such as cancer, diabetes or autoimmune disorders are multifactorial (Cho and Gregersen, 2011; Pearson et al., 2003; Wu et al., 2018). Thus, molecular interactions, as defined by the correlation across different omics and reported here, may substantiate previous discoveries related to any single molecule (gene, protein metabolite) or the interactions between them, defined by e.g. pQTLs or mQTLs. We used COmics to provide an overview on these molecular interactions and thus further insight into complex disease. We focused on T2D **(BOX1)**; cancer **(BOX2)**; and cardiovascular diseases **(BOX3)**, for which we prepared linked to a molecular network located in COmics server along with the brief description in the BOX, designated for each disease. By giving those examples we would like to encourage you to utilize COmics server (http://comics.metabolomix.com) as a resource to explore molecular interactions related to physiological processes but also to complex diseases.

## DISCUSSION

Understanding platform correlation and complementarity is central to working with large genetic and epidemiological meta-analyses, evaluation of data integration options, as well as exploring additional information in already measured datasets. By investigating MBH between omics platforms covering overlapping molecules we found that metabolomics and glycomics platforms are characterized by multiple MBH linked identical molecules, which underscores quality of the measurements conducted and suggest feasibility of data integration. For the two affinity-based platforms used for proteomics, approximately 20% of common protein targets were not detected by the MBH and suggest that integration of the measurement approaches may be challenging and require special attention, especially for the molecules which were not identified by MBH. Our observations are in line with previous study assessing proteomics methods in multiple cohorts (Raffield et al., 2020), and could be linked with e.g. different sensitivity of aptamer/antibody towards the protein target.

We previously showed that further insight into the complex lipid structures such as phosphatidylcholines is achievable by combining data sets which integrate measurements of various lipids from different platforms (Quell et al., 2019). Here, using MBH we replicated that finding as well as provided further insight into composition of triglycerides, which is of relevance for future lipidomics studies.

Integration of multiomics layers may contribute to identification of processes relevant to human physiology and pathophysiology and thus to the integrative representation of the Molecular Human. GWAS or EWAS with intermediate phenotypes e.g. miRNA, protein, glycan or metabolite have shown potential in providing insight into human physiology and complex diseases in the past (Chen et al., 2012; Dai et al., 2021; Hasin et al., 2017; Karczewski and Snyder, 2018; Sailani et al., 2020; Suhre et al., 2010). The integration of additional multiomics layer resulted in identification of processes relevant for human physiology and pathophysiology including instances mirroring biochemical reactions and metabolism of the components involved, as well as molecular interactions previously identified by multiomics GWAS, EWAS and TWAS. We reproduced a plethora of literature reported hits, provingrobustness of our approach, as well as identified new associations shedding new light on a range of biological processes.

Finally, we provided showcase examples for molecular networks relevant to complex disease such as T2D, cancer and cardiovascular disease. Those revealed previously unreported molecular associations beyond molecular events known to be related to the conditions investigated and provide promising candidate mechanisms to explore in future work in the study of the respective disease.

Our study has strengths and weaknesses. Participants were enrolled continuously on an availability basis (i.e. without selection for diabetes state, age, sex, BMI or ethnicity) at the dermatology department of the major public hospital in Doha, Qatar, using identical collection kits and protocols, in order to avoid any possible batch effects between cases and controls. The large diversity of the QMDiab participants provides access to a wide range of individuals from a broad range of lifestyles, which may be advantageous when investigating correlations between omics layers, as the signal to noise ratio may increase. Similarly, the fact that study participants were not fasting implies further biological variation in the data, which may strengthen correlation signals that are related to processes confounded by fasting when case-control studies are conducted.

Taken together, we have drawn an image of the Molecular Human by providing a comprehensive description of biologically relevant molecular interactions in the human body based on the integrated data generated by 18 technologically diverse platforms in human samples obtained from 391 subjects. We provide access to this resources via the COmics web-server and Github. Our study describes the complementarity of various omics layers, demonstrates the capacity for integrated omics data to mirror biological processes, and is further setting the stage for future studies which may utilize this resource to understand the molecular network surrounding molecules of interest, with the potential of linking it to the disease end points.

## METHODS

### Cohort characteristics

The subjects were enrolled in the framework of the Qatar Metabolomics Study on Diabetes (QMDiab), a cross-sectional diabetes case-control study at the Dermatology Department of HMC in Doha, Qatar as previously described (Mook-Kanamori et al., 2014). The study was approved by the Institutional Review Boards of HMC and Weill Cornell Medicine, Qatar (WCM-Q) (research protocol #11131/11). Written informed consent was obtained from all participants. The study enrolled 391 participants with at least one omics phenotype and includes 17 additional subjects that were not a part of Mook-Kanamori et al. The cohort consists of 198 females and 193 males. The average participants age was 46.5 years (s.d. = 12.9) and the average BMI was 29.7 kg/m^2^ (s.d. = 6.0). This cohort includes 195 participants with T2D and 196 without T2D.

### Sample collection

Non-fasting blood, saliva and urine were collected according with standard protocols as previously described (Mook-Kanamori et al., 2014). Blood was collected using EDTA, Heparin, citrate and PAXgene Blood RNA tubes. Blood collected in EDTA and Heparin was centrifuged at 2,500 g for 10 min, plasma was collected aliquoted and stored at -80°C until analysis. The blood collected into PAXgene Blood RNA tubes was centrifuged for 10 min at 4,000 g. The supernatant was removed, and the pellet was used for the RNA extraction. The saliva was collected using Salivette system (Salivette®, SARSTEDT AG & Co. KG) according with manufacturer’s protocol. Collected saliva samples were centrifuged at 2000 × g for 2 minutes, aliquoted and stored at -80°C until analysis. The urine was collected into the URINE CAPS mixed transferred into the falcon tube, centrifuged at 2,500 g for 10 min, aliquoted and stored at -80°C.

### Deep molecular phenotyping

The obtained samples were submitted for deep molecular phenotyping which utilized clinical chemistry parameters along with omics measurements across 18 technically diverse platforms. We determined: 41 clinical chemistry parameters (CLIN); genotype data of 1,221,345 variants (DNA) ; 450k DNA methylation sites (MET); 4) 57,942 transcriptomic traits, including 57,773 RNA transcripts (RNA) using RNA-sequencing (Illumina, 20M reads), and 169 microRNA profiles (miRNA) with multiplex qPCR (Exicon); 1,313 blood circulating proteins using two different technologies 1129 proteins (SOMA) from aptamer-based technology (SomaLogic) and 184 proteins (OLINK) from high-multiplex immunoassays (Olink); 274 glycan traits including 36 total plasma N-glycosylation (PGP) using HILIC-UPLC and 60 IgG-glycopepdides (IgG) deploying LC-MS, both profiled at Genos Ltd. as well as 178 IgA and IgG-glycopeptides (IgA) measured with LC-MS in Wuhrer lab; 225 plasma lipoproteins (BRAIN) quantified with ^1^H NMR (Nightingale), 1494 lipids including 1,331 plasma lipids (LD) quantified using Lipidyzer deploying LC-MS system (Metabolon), and 163 plasma lipids and other metabolites (BM) quantified with FIA-MS (Biocrates p150 kit); 3,415 metabolic traits profiled with different approaches and matrixes including 1,104 plasma metabolites (HDF) determined with HILIC-MS and UPLC-MS on HD4 platform (Metabolon), 2,251 metabolites (758 in plasma (PM), 602 in saliva (SM) 891 in urine (UM),) measured using GC-MS and UPLC-MS on HD2 platform (Metabolon), and 60 urine lipids (CM) quantified with ^1^H NMR deploying Chenomx (University Greifswald). For the cross-platform analyses we limited the RNA profiles to 1239 transcripts, which were also assayed by SOMA and OLINK platforms.

### Clinical chemistry data

The obtained blood samples were analyzed within four hours of blood collection at the Department of Laboratory Medicine and Pathology of HMC with the Cobas® 6000 (Roche Diagnostics, Basel, Switzerland).

### Transcriptomics (RNA-seq)

The obtained pellets from the PAXgene Blood RNA tubes were used for the isolation of total RNA with PAXgene Blood miRNA Kit (Qiagene). In brief, the obtained pellets were mixed with RNaze-free water, and vortexed until the pellets dissolved. The samples were centrifuged for 10 min at 4000 x g and pellet was formed. The supernatant was removed and 350 μL of BM1 buffer provided with the kit was added into the pellet. The samples were vortexed until the pellet dissolved, and mixed with 300 μL of BM2 buffer as well as 40 μL of proteinase K, provided with the kit. The samples were incubated for 10 min. at 55°C under constant shaking followed by transfer onto the PAXgene shredder spin column placed in a processing tube. The samples were centrifuge for 3min at 15,000 x g and the supernatant was placed into the fresh tube, mixed with 700 μL of 100% isopropanol and transferred onto PAXgene RNA spin column. The samples were centrifuged for 1 min at 15,000 x g the flow-throw was removed, and 350 μL of BM3 buffer was placed onto PAXgene RNA spin column. The samples were centrifuged for 15 sec. at 15,000 x g, the PAXgene RNA spin column was placed in the fresh collection tube and 80 μL of RDD buffer containing DNase-I was placed onto PAXgene RNA spin column followed by 15 min. incubation at room temperature. 350 μL of BM3 buffer was placed onto PAXgene RNA, the samples were centrifuged for 15 sec. at 15,000 x g, the flow-throw was removed, and 500 μL of BM4 buffer was added. The samples were centrifuged for 15 sec. at 15,000 x g, the flow-throw was removed, and additional 500 μL of BM4 buffer was added. After centrifugation for 2 min. at 15,000 x g, the PAXgene RNA spin column was placed into the fresh collection tube, and the samples were eluted from the column with 80uL of BR5 buffer. The obtained eluent was incubated for 5 min at 60°C, and afterwards chilled on ice. The integrity and quantity of the isolated RNA was measured using Qubit RNA HS Assay Kit (high sensitivity, 5 to 100 ng quantification range) Assay Kit and Qubit 3.0 fluorometer (Life Technologies) according to the manufacturer’s protocol. The samples were kept at -80 °C until measurements.

The samples containing total RNA (400 ng) were submitted to the Genomics Core at WCMQ for the RNA-sequencing. The total RNA was depleted of rRNA and Globin using the NEBNext rRNA & Globin Depletion Kit for Human/Mouse/Rat (New England BioLabs, Ipswich, MA). The depleted RNA was used to generate strand-specific libraries with BIOO NEXTflex Rapid Directional RNA-Seq Kit (Bioo-Scientific, Austin, TX). Library quality and quantity were analyzed with the Bioanalyzer 2100 (Agilent, Santa Clara, CA) on a High Sensitivity DNA chip. 10 libraries were then pooled in equimolar ratios and paired-end sequenced at 75bp on one lane of an Illumina HiSeq 4000 (Illumina, San Diego, CA). Total of 57,773 RNA transcripts were measured in 320 subjects.

### microRNA quantification

#### RNA extraction

The miRNAs were isolated from 200μL EDTA-plasma sample using the miRNeasy serum/plasma kit (Qiagen) following the manufacturer’s instructions. Briefly, the samples were lyzed using QIAzol Lysis Reagent and spiked with 3.5 μl miRNeasy Serum/Plasma Spike-In Control included in the kit. The chloroform was added, samples were mixed and the centrifuged. The obtained after centrifugation upper aqueous phase was transferred into the fresh tube, mixed with 1.5 volume of 100% ethanol, and transferred into an RNeasy MinElute spin column in a 2 ml collection tube, provided in the kit. The samples were centrifuged, the flow-throw was removed, and RWT buffer provided with the kit was added onto the RNeasy MinElute spin column. The samples were centrifuged, the flow-throw was discarded, RPE buffer, provided with the kit, was added onto the RNeasy MinElute spin column. The samples were centrifuged and flow-throw was removed. The 80% ethanol prepared in RNaze-free water was placed onto the MinElute spin column, the samples were centrifuged until the spin column membrane dried. The MinElute spin column was placed in fresh collection tube and the total RNA including miRNA was eluted with 14 μl RNase-free water.

#### miRNA profiling

Prior the profiling, the isolated RNA samples were reverse transcribed to cDNA using the Exiqon Universal cDNA Synthesis Kit II (Exiqon Inc., MA, USA) according with the manufacturer instruction. Briefly, 2 μL of total RNA (5 ng/μL) were used for cDNA synthesis. All processes were conducted in 384 well plate format. The quality and integrity of the synthesized cDNA was assessed using the miRNA QC PCR Panel (V4.M; Exiqon Inc.). Obtained cDNA was 50-fold diluted and mixed with 2x Exilent SYBR Green master mix (Exiqon Inc.), and ROX reference dye (4 μl/2 ml) (Thermo Fisher Scientific, MA, USA). The samples were loaded onto human serum/plasma focus miRNA PCR panels, and quantitative real-time PCR was performed using the QuantStudio 12 K Flex real-time PCR System (Applied Biosystems, CA, USA). The PCR data were processed using Exiqon GenEx qPCR analysis software (version 6). The inter-plate calibration was performed using the mean value of UniSp3 interplate calibrator. The samples with a high degree of hemolysis were identified after monitoring of calculated ΔC_t_ between hsa-miR-23a-3p and hsa-miR-451a. The samples with ΔC_t_ >7 were removed from the analysis. Only microRNA assays with C_t_ ≤ 35, expressed in at least 60% of the samples were counted and the remaining samples were removed from the analysis. The global average of all expressed microRNAs with C_t_ <35 was used to normalize individual assays. Total of 169 miRNAs were profiled in 339 subjects.

### Proteomics measurements using SOMAscan technology

The EDTA-plasma samples were used for proteomics analysis based on SOMAscan assay (version 1.1) technology, which was conducted at the WCM-Q Proteomics Core (Suhre et al., 2017). The method employed protein-capture by *Slow Offrate Modified Aptamers* (SOMAmer) (Gold et al., 2010). Briefly, undepleated EDTA-plasma was diluted and the following assay steps were performed: 1) Binding: analytes and SOMAmers, carrying a biotin moiety via a photocleavable linker were equilibrated; 2) *Catch I*: analyte/SOMAmer complexes were immobilized on streptavidin-support, followed by washing steps to remove proteins not stably interacting with SOMAmers; 3) *Cleave*: release of analyte/SOMAmer complexes from streptavidin beads through exposure to long-wave ultraviolet light resulting in linker cleavage; 4) *Catch II*: biotinylation of proteins in analyte/SOMAmer complexes and subsequent repeated immobilization on streptavidin support followed by washing steps to select against non-specific analyte/SOMAmer complexes; 5) *Elution*: denaturation of analyte/SOMAmer complexes and SOMAmer release; 6) *Quantification*: hybridization to custom arrays of SOMAmer-complementary oligonucleotides. The primary data were submitted to Somalogic for normalization of raw intensities, across-batch calibration and steps of quality control. In total 1129 molecules were quantified in 356 samples.

### Proteomics measurements using Olink technology

Heparin-plasma samples were used for the proteomics measurements based on the Olink® technology (Olink Proteomics AB, Uppsala, Sweden) at the WCM-Q Proteomics Core. The technology is based on a proximity extension assay (PEA) (Assarsson et al., 2014), and enables for simultaneous analysis of 92 analytes in 1 μL of sample. We used two different Olink® panels, namely Cardiometabolic and Metabolism, for measurements of 184 unique proteins. The samples were processed along with 8 control samples according to the manufacturer’s protocol using the following steps: 1) Immunoassay: the sample was mixed and incubated with 92 supplier-provided optimized antibody pairs labeled individuallywith oligonucletotides (PEA probes). Pair coupled oligonucleotides carry unique annealing sites that allows specific hybridization of matching probes; 2) Extension: Target binding by antibody pairs brings the corresponding probe oligonucleotides in close proximity and allows for hybridization. Hybridized templates are extended by DNA polymerase, which generates a DNA template for amplification; 3) Preamplification: Universal primers enable parallel preamplification of all 92 DNA templates by PCR; 4) Detection: The resulting DNA sequence is subsequently detected and quantified using a microfluidic real-time PCR instrument (Biomark HD, Fluidigm, South San Francisco, CA, USA). The data obtained were normalized using an internal extension control and an inter-plate control, to adjust for intra- and inter-run variation. In total 184 proteins were quantified in 328 samples.

### Total Plasma N-Glycosylation (Genos platform)

#### Sample processing

he EDTA-plasma samples were analyzed by Genos Ltd. (Zagreb, Croatia) using ultra-performance liquid chromatography (UPLC) glycoprofiling as previously described (Suhre et al., 2019; Trbojević Akmačić et al., 2015). Briefly, the sample processing for total plasma N-glycosylation measurements was conducted in 96-well plate format out of 10 μL of plasma sample in following steps: 1) Release of N-glycans from plasma proteins: The plasma proteins were denaturated with 20 μl of sodium dodecyl sulfate (SDS) 2% (w/v) (Invitrogen, USA) for 10 min at 65°C, followed by cooling to room temperature for 30 min, and mixing with 10 μl of 4% (v/v) Igepal-CA630 (Sigma-Aldrich, USA) under constant shaking for 15 min. N-glycans were released after incubation of samples with enzyme, N-glycosidase-F (1.2 U of PNGase F (Promega, USA)) overnight at 37°C; 2) Fluorescent labeling of released plasma glycans: The obtained N-glycans were mixed with freshly prepared labeling mixture containing (70 : 30 v/v) 2-aminobenzamide and 2-picoline borane in dimethylsulfoxide (Sigma-Aldrich) and glacial acetic acid (Merck, Germany) for 15 min followed by 2 h incubation at 65°C; 3) Cleaning and elution of labeled N-glycans: The excess free label and reducing agent were removed from the samples using hydrophilic interaction liquid chromatography solid-phase extraction (HILIC-SPE). The samples were loaded into the wells of 0.2 μm 96-well GHP filter-plate (Pall Corporation, USA), which was used as stationary phase, and were washed 5 times with cold 96% acetonitrile (ACN). Glycans were eluted with 2 × 90 μL of ultrapure water under constant shaking for 15 min at room temperature. The eluates were combined and stored at -20 °C until use.

#### Sample measurements

Total plasma *N*-glycans were measured using HILIC-UPLC as previously described (Trbojević Akmačić et al., 2015). Briefly, the labeled *N*-glycans were gradient eluted from Waters BEH Glycan chromatography column (Waters UPLC BEH particles 2.1⍰× ⍰150⍰mm, 1.7⍰μm) using 100 mM ammonium formate at pH 4.4, and ACN. The flow rate was 0.56 ml/min in a 23 min of the analytical run. The fluorescence was measured at 420 nm with excitation at 330 nm using Waters Acquity UPLC H-class system consisting of a fluorescence (FLR) detector set with 250 nm excitation and 428 nm emission wavelengths.

The data processing was performed using an automatic processing method enabling to obtain chromatograms separated into 39 peaks. The data was further quantified and annotated into 36 primary glycan traits (Trbojević Akmačić et al., 2015). All N-glycans have core sugar sequence consisting of two N-acetylglucosamines (GlcNAc) and three mannose residues; F indicates a core fucose α1–6 linked to the inner GlcNAc; Ax indicates the number of antennas (GlcNAc) on trimannosyl core; Gx indicates the number of β1–4 linked galactoses on antenna; G1 indicates that the galactose is on the antenna of the α1–6 mannose; Sx indicates the number (x) of sialic acids linked to galactose. In total 36 total plasma *N*-glycans were measured in 345 subjects.

### IgG glycosylation (Genos platform)

#### Sample processing

The IgG isolation and measurements were conducted by Genos, Ltd as previously described (Pučić et al., 2011; Sharapov et al., 2019). Briefly, the sample processing for plasma IgG-glycosylation measurements was conducted in the following steps: 1) Preparation of protein G monolithic plates: the 96-well protein G monolithic plate (BIA Separations, Ajdovščina, Slovenia) was washed with 10 column volumes of ultrapure water, 10 column volumes of binding buffer (1 x PBS), and 5 column volumes of 0.1 M formic acid (pH 2.5). The protein G plate was equilibrated with 10 column volumes of 10 x binding buffer and 20 column volumes of 1 x binding buffer; 2) Isolation of IgG from human plasma: For the IgG isolation the protein G monolithic plate was used. The IgG were obtained from 70 - 100 μl of plasma. The samples were diluted 10 times with binding buffer and filtered through *GHP AcroPrep* 96-well filter plates. The samples were applied onto the protein G monolithic plates and instantly washed three times with PBS to remove the unbound proteins; 3) Elution of IgGs: The IgG were eluted from the protein G monoliths into 96-well plate with 5 column volumes of 0.1 M formic acid (Merck, Germany) followed by immediate neutralization with 1 M ammonium bicarbonate (Merck, Germany) (Menni et al., 2013); 4) IgG digestion and purification: Aliquots of 40 μl from the obtained samples, containing isolated IgG, were used for further processing. The samples were incubated with 2% SDS [20 μL (w/v)] for 10 min at 60°C, and followed by overnight incubation with 200 ng trypsin at 37°C. The obtained IgG tryptic glycopeptides samples were purified by reverse phase solid phase extraction using Chromabond C18 beads applied to each well of an OF1100 96-well polypropylene filter plate. The beads were activated with 80% ACN containing 0.1% trifluoroacetic acid (TFA); 5) IgG elution: The tryptic digests were diluted 10 times in 0.1% TFA, loaded onto the C18 beads in vacuum manifold and washed 3 times with 0.1% TFA. IgG glycopeptides were eluted into an PCR 96 well plate with 120 μl of 20% ACN containing 0.1% TFA by 5 min centrifugation at 15-105 x g. Eluates containing glycopeptides were dried by vacuum centrifugation and - 20°C until analysis by MS.

#### Sample measurement

Purified tryptic IgG glycopeptides were analyzed as previously described (Pučić et al., 2011). For the separation and measurements nanoACQUITY UPLC system (Waters, Milford Massachusetts, USA), consisting of binary pump, auxillary pump, autosampler maintained at 10 °C and column oven compartment set at 30 °C coupled to and the Bruker Compact Q-TOF-MS were used. 9 μL of purified IgG glycopeptides sample was applied to a Thermo Scientific PepMap 100 C8 (5 mm × 300 μmi.d., 5 μm) SPE trap column. After sample loading the trap column was switched in-line with the gradient and C18 nano-LC column (150 mm × 100 μm i.d., 2.7 μm HALO fused core particles; Advanced Materials Technology, Wilmington, Delaware, USA) for 9.5 min while sample elution took place. IgG glycopeptides were reconstituted in 20 μl MQ water before nano-LC-ESI-MS analysis. Separation was achieved at 1 ml/min using the following gradient of mobile phase A and mobile phase B (80 % ACN and 20 % 0.1 % TFA): 0.5 min 12 % B, 0.5 - 4 min 12 % B - 17 % B, 4 - 5 min 17 % B. The column outlet tubing was directly applied as sprayer needle. Quadrupole and collision energy was set at 4 eV. Spectra were recorded from m/z 600 to 1900 with 2 averaged scans at a frequency of 0.5 Hz. Per sample the total analysis time was 15 min.

The nanoACQUITY UPLC system and the Bruker Compact Q-TOF-MS were operated under HyStar software version 3.2.

Glycan data was first normalized (total area normalization) and then batch corrected using Combat. Batch correction was performed on the log-transformed normalized data. After batch correction, the data was inverse transformed so all values were between 0 and 100. Finally the data was z-scored. Glycan structural features are given in terms of number of galactoses (G0, G1 and G2), fucose (F), bisecting N-acetylglucosamine (N) and N-acetylneuraminic acid (S). Total of 60 IgGs were measured in 341 samples.

### IgA and IgG glycosylation (Univ. Leiden platform)

#### Sample processing

The purification, separation and measurements of IgA and IgG was conducted at Leiden University Medical Center as previously described (Dotz et al., 2021; Momcilovic et al., 2020). Briefly, 2 μL and 5 μL of plasma was used for IgG and IgA analysis, respectively. The samples were diluted with PBS to obtain final volume of 200 μL. The samples purification was conducted in duplicate on separate plates using affinity bead chromatography. The samples designated for IgG analysis were purified using 15 μL/well of Protein G Sepharose 4 Fast Flow beads (GE Healthcare) on an Orochem filter plate, followed by three washing steps with PBS. The samples designated for IgA analysis were purified using 2 μL/well of CaptureSelect IgA Affinity Matrix beads (Thermo Fisher Scientific). The plates were incubated for 1 h under constant shaking.

The samples were washed three times with PBS followed by three additional washes with purified water using vacuum manifold. The IgGs and IgAs were eluted from the beads using 100 mM formic acid under constant shaking for 10 min, followed by 1 min centrifugation at 100 x g. The obtained eluates were dried for 2 h at 60°C in a vacuum centrifuge.

The samples designated for IgG analysis, were resolubilized by addition of ammonium bicarbonate (50 mM) under constant shaking for 5 min. The samples were digested by overnight incubation with tosyl phenylalanyl chloromethyl ketone (TPCK)-treated trypsin at 37°C.

The samples designated for IgA analysis were reduced and alkylated prior to digestion to obtain peptides covering all glycosylation sites. The samples resolubilization was conducted with ammonium bicarbonate (30 mM) containing 12.5% of acetonitrile under constant shaking for 5 min. The samples were mixed with dithiothreitol (35 mM) and incubated for 5 min at room temperature followed by additional incubation for 30 min at 60°C. The samples were cooled to room temperature, mixed with iodoacetamide (125 mM), incubated in the dark under shaking for 30 min and mixed with dithiothreitol (100 mM) to quench the iodoacetamide. The samples were digested with TPCK-treated trypsin by the incubation over night at 37°C.

#### Sample measurement

The samples designated for IgG and IgA were measured at different days. The sample separation and measurements were conducted on Ultimate 3000 RSLCnano system (Dionex/Thermo Scientific) equipped with an Acclaim PepMap 100 trap column (particle size 5 μm, pore size 100 Å, 100 μm × 20 mm) and an Acclaim PepMap C18 nano analytical column (particle size 2 μm, pore size 100 Å, 75 μm × 150 mm) coupled to a quadrupole-TOF-MS (Impact HD; Bruker Daltonics). 250 μL of sample was injected into the flow (25 μL/min) of aqueous solvent and was trapped on the trap column (Dionex Acclaim PepMap100 C18, 5 mm × 300 μm; Thermo Fisher Scientific, Breda, The Netherlands). The analytes were eluted on the analytical column (Ascentis Express C18 nanoLC column, 50 mm × 75 μm, 2.7 μm fused core particles; Supelco, Bellefonte, PA) under flow rate of 0.9 μL/min and separated in linear gradient from 3% to 30% solvent containing 95% (v/v) ACN. The samples were measured in positive-ion mode using a CaptiveSprayer (Bruker Daltonics) electrospray source at 1300 V. The mass spectra were acquired with a frequency of 1 Hz and the MS ion detection window was set at mass-to-charge ratio (m/z) 550–1800. Fragmentation spectra were recorded with a detection window of m/z 50–2800.

Obtained LC-MS data were examined according with pipeline developed by Manfred Wuhrer lab as previously described (Dotz et al., 2021; Momcilovic et al., 2020). In total 178 molecules including IgGs and IgAs were measured in 344 samples.

### Untargeted metabolomics – Metabolon HD2 platform

The EDTA-plasma, saliva, and urine samples were used for untargeted metabolic profiling as we previously described (Mook-Kanamori et al., 2014; Yousri et al., 2015). The measurements were conducted at Metabolon Inc, deploying HD2 platform based on ultra-high-performance liquid chromatography-mass spectrometry (UPLC-MS) and gas chromatography-mass spectrometry (GC-MS) technology (Evans et al., 2009). In brief, the sample was mixed with the recovery standards prior to the extraction for quality control (QC) purposes. The resulting sample extract was divided into aliquots designated for the analysis using the following: 1) UPLC-MS/MS with positive ion mode electrospray ionization (ESI); 2) UPLC-MS/MS with negative ion mode ESI; 3) hydrophilic interaction chromatography (HILIC)/UPLC-MS/MS; 4) GC-MS. The sample extract was dried under nitrogen flow and reconstituted in solvents compatible with each of the four analytical methods.

Three out of the four sample aliquots were designated for LC-MS measurements and were reconstituted in acidic or basic solvents. The first sample aliquot was reconstituted in acidic conditions, and gradient eluted from a C18 column (Waters UPLC BEH C18-2.1×100 mm, 1.7 μm) with water and methanol containing 0.1% formic acid (FA). The second sample aliquot was reconstituted in basic solvent, and gradient eluted from C18 column (Waters UPLC BEH C18-2.1×100 mm, 1.7 μm) with water and methanol containing 6.5mM ammonium bicarbonate. The third aliquot was gradient eluted from a HILIC column (Waters UPLC BEH Amide 2.1×150 mm, 1.7 μm) using water and acetonitrile with 10mM ammonium formate. The flow rate was 350 μL/min, and the sample injection volume was 5 μL.

The separation and measurements of the sample aliquots designated for LC-MS were performed on Waters ACQUITY UPLC in-line to Thermo Scientific Q-Exactive high resolution/accurate mass spectrometer interfaced with a heated electrospray ionization (HESI-II) source and an Orbitrap mass analyzer. In the MS analysis, the scan range varied between methods but fell within the range of 70-1000 m/z.

The remaining fourth sample aliquot was designated for GC-MS measurements. The sample aliquot was derivatized with N,O-Bis(trimethylsilyl)trifluoroacetamide (BSTFA) followed by drying under nitrogen flow. Separation was conducted under temperature ramp from 60 – 340°C over a period of 17.5 min, using a 5% diphenyl / 95% dimethyl polysiloxane fused silica column (20 m × 0.18 mm ID; 0.18 um film thickness) and helium at flow rate of 1 ml/min as the carrier gas. The measurements were performed on a Thermo-Finnigan Trace DSQ fast-scanning single-quadrupole mass spectrometer using electron impact ionization (EI), and the MS scan range was from 50-750 m/z.

The number of measured metabolites in given sample matrix was following: 758 metabolites in 358 EDTA-plasma samples, 602 metabolites in 283 saliva samples, and 891 metabolites in 360 urine samples.

### Untargeted metabolomics – Metabolon HD4 platform

The EDTA-plasma samples were used to conduct metabolic profiling at Metabolon Inc on technologically advanced, in comparison with HD2, HD4 platform enabling for increased sensitivity and accurate detection of more metabolites. The main technical difference between HD2 and HD4 platforms was replacement of GC-MS with hydrophilic interaction chromatography (HILIC) method. The method was described in great detail previously (Evans, 2014). In brief, sample processing was conducted as we described in “*Untargeted profiling - HD2 platform: LC-MS and GC-MS*” section, except of the sample dedicated for GC-MS measurement. This sample aliquot instead was gradient eluted from a HILIC column (Waters UPLC BEH Amide 2.1⍰×1⍰50⍰mm, 1.7⍰μm) using water and acetonitrile with 10 mM ammonium formate at pH 10.8. The measurements were conducted using Waters ACQUITY ultra-performance liquid chromatography (UPLC) and a Thermo Scientific Q-Exactive high-resolution/accurate mass spectrometer interfaced with a heated electrospray ionization (HESI-II) source and Orbitrap mass analyzer (Evans, 2014). Total of 1,104 metabolites were measured in 309 plasma samples.

### Targeted metabolomics – Biocrates p150 platform

The EDTA-plasma samples were used for targeted metabolomics analysis. The samples were measured at the Metabolomics Platform of the Helmholtz Center Munich using AbsoluteIDQ™ kit p150 (Biocrates Life Science AG, Innsbruck, Austria) as previously described (Illig et al., 2010; Römisch-Margl et al., 2012). The AbsoluteIDQ p150 kit assay enables the quantification of up to 163 molecules, predominantly lipids including phospatidylcholines (PCs), lysoPCs, sphingomyelines (SM), and acylcarnitines (AC), as well as amino acids. Total of 10 μL of plasma was used to conduct the assay. The samples were applied on the assay kit 96-well plate consisting of filters with internal standards and were dried under a nitrogen stream at room temperature (RT). The samples were derivatized with a reagent containing 5% phenylisothiocyanate (PITC), dryed under a nitrogen stream at RT, and extracted with 300 μL of 5 mM ammonium acetate in methanol. Next, the samples were filtered by centrifugation, the resulted flow-through was diluted 1:6 with running solvent and placed into fresh deep-well plate. The plate was covered with the silicone mat, and mixed. Sample handling was performed by a Hamilton Microlab STARTM robot (Hamilton Bonaduz AG, Bonaduz, Switzerland) and a Ultravap nitrogen evaporator (Porvair Sciences, Leatherhead, U.K.), beside standard laboratory equipment. Metabolites were measured in positive and negative multiple reaction monitoring (MRM) scan mode by direct infusion to an API 4000 triple quadrupole system (SCIEX Deutschland GmbH, Darmstadt, Germany) equipped with a 1200 Series HPLC (Agilent Technologies Deutschland GmbH, Böblingen, Germany) and a HTC PAL auto sampler (CTC Analytics, Zwingen, Switzerland) controlled by the software Analyst 1.6.2. The metabolite concentrations were calculated using internal standards and the MetIDQ software provided with AbsoluteIDQ™ kit, and are reported in μmol/L. For the lipid molecules including PC, lysoPC, SM, and AC, measured with AbsoluteIDQ™ kit the information on the sum of the carbons of the fatty acid chains is provided but not the fatty acid chain actual composition. For example, PC.aa.36.1 describes phosphatidylcholine (PC) where two glycerol residues are bound in diacyl (aa) binding into the fatty acid moiety; the sum of carbons of both fatty acid chains is 36, and there is one double bound (.1). Total of 163 metabolites were quantified in 356 samples.

### Lipidomics – Lipidyzer platform

The EDTA-plasma samples were used for in depth profiling of lipids, which was conducted at Metabolon Inc. deploying Lipidyzer™ platform of AB Sciex Pte technology as previously described (Löfgren et al., 2012; Quell et al., 2019). In brief, the samples were extracted in the presence of internal standards using butanol:methanol (BUME) mixture (3:1) followed by two-phase extraction into 300 μl heptane:ethyl acetate (3:1) using 300 μl 1% acetic acid as buffer. The obtained extracts were dried under nitrogen flow and reconstituted in ammonium acetate dichloromethane:methanol. The samples were analyzed in both positive and negative mode electrospray using Sciex SelexIon-5500 QTRAP. The molecules were detected in MRM mode with a total of more than 1,100 MRMs. Individual lipid species were quantified by the ratio of the signal intensity of each target compound to that of its assigned internal standard, followed by the multiplication of the concentration of internal standard added to the sample. Lipid class concentrations were calculated from the sum of all molecular species within a class, and fatty acid compositions were determined by calculating the proportion of each class comprised by individual fatty acids. Total of 1,331 lipids were measured in 324 samples.

### NMR metabolomics - urine

1H-NMR spectra analysis of urine samples was conducted at Institute of Clinical Chemistry and Laboratory Medicine, University of Greifswald, Germany as previously described (Budde et al., 2016; Zaghlool et al., 2018).In brief, Bruker DRX-400 NMR spectrometer (Bruker BioSpin GmbH, Rheinstetten, Germany) operating at 400.13 MHz ^1^H frequency equipped with 4 mm selective inverse flow probe (FISEI, 120 μL active volume) was used to record the spectra. 500 μL of sample volume was delivered via automatic flow injection. The sample acquisition temperature was 300 K. A standard one-dimensional ^1^H NMR pulse sequence with suppression of water peak (NOESYPRESAT) was used (Budde et al., 2016). The free induction decays (FIDs) were collected into data points using spectral width of 20.689 ppm. The FIDs were multiplied by an exponential function corresponding to 0.3 HZ line-boarding prior to Fourier-transformed (FT). For the assessment of the spectra quality, the line width and signal-to-noise ration of Trimethylsilylpropanoic acid (TSP) signal, used as a reference, was analyzed. Additionally, quality control was carried out by analyzing the standard error of creatinine concentration and the potential variability of selected signals. The obtained spectra were processed within TOPSPIN 1.3 (Bruker BioSpin GmbH) and the metabolites annotation and quantification was conducted in semi-automated manner by spectral pattern matching using Chnomx NMR suit 7.0 (Chenomx Inc.). Total of 60 lipid molecules were measured in 353 samples.

### NMR metabolomics - plasma

The EDTA-plasma (300 μL) was used for metabolite quantification by a high-throughput NMR metabolomics platform (Nightingale Ltd, Helsinki, Finland) (Soininen et al., 2009; Zaghlool et al., 2018). The sample preparation was conducted automatically using Gilson Liquid Handler 215. Each sample after brief centrifugation was transferred to SampleJet NMR tubes and mixed with 300 μL of sodium phosphate containing 0.08% of TSP. The measurements were conducted on Bruker AVANCE III 500 MHz and Bruker AVANCE III HD 600 MHz spectrometers. The lipoprotein (LIPO) and low-molecular-weight metabolites (LMWM) were measured in the samples using either 500 MHz or 600 MHz spectrometers. The same samples were further extracted with multiple extraction steps as previously detailed (Soininen et al., 2015). The extracted lipid (LIPID) data was evaluated in full automation with the 600 MHz instrument using a standard parameter set (Soininen et al., 2015). The FT and automated phasing of NMR spectra was conducted followed by automated spectral processing and quality control steps (Soininen et al., 2009). The subclasses for the lipoproteins are categorized according to size following this classification: chylomicrons and extremely large VLDL particles (average particle diameter at least 75 nm); five VLDL subclasses—very large VLDL (average particle diameter of 64.0 nm), large VLDL (53.6 nm), medium VLDL (44.5 nm), small VLDL (36.8 nm) and very small VLDL (31.3 nm); intermediate-density lipoprotein (IDL; 28.6 nm); three LDL subclasses—large LDL (25.5 nm), medium LDL (23.0 nm) and small LDL (18.7 nm); and four HDL subclasses—very large HDL (14.3 nm), large HDL (12.1 nm), medium HDL (10.9 nm) and small HDL (8.7 nm). Total of 225 molecules were measured in 350 samples.

### Statistical data analysis

All statistical analyses were conducted using R (version 4.1.0 and above) and Rstudio (version 1.4.1717 and above). If not otherwise stated, the omics data was converted “as-received” into R Summarized Experiment format, representing processed final data. The saliva metabolomics data has been further normalized by saliva osmolality, and the urine metabolomics data has been normalized by urine creatinine.

### Cross-platform correlations (omicsMBHs)

Spearman correlation coefficients between unscaled raw omics data were computed and mutual best hits were identified. Platform-pairwise Bonferroni significance cutoffs (p < 0.05 / (n_PLTA1_ * n_PLAT2_ / 2)) were used.

### Within-trait partial correlations (GGMs)

Partial correlations within platforms were computed as follows: Saliva and urine metabolites were normalized by saliva osmolality and urine creatinine obtained from the respective platform, respectively. The omicsdata was then inverse-normal scaled. Metabolites and then samples with more than 50% missing values were removed. Association statistics and residuals were then computed using the linear model “lm(OMICS ∼ AGE + SEX + BMI + DIAB + genoPC1 + genoPC2 + genoPC3 + somaPC1 + somaPC2 + somaPC3)”. Missing values were imputed using the K-nearest-neighbors method (Do et al., 2018). Partial correlation coefficients were computed using the pcor function from the R-package GeneNet (version 1.2.15). Platform-wise Bonferroni significant correlations (p-value < 0.05 / (NPLAT*(NPLAT-1)/2)), where NPLAT represents the number of traits measured on the respective platform, were retained.

### Association between DNA – RNA – METH (eQTLs, eQTMs, meQTLs)

Genetic variants (SNPs) were coded 0, 1, 2 for major allele homozygotes, heterozygotes, and minor allele homozygotes, respectively. Expression data was log-scaled, with all values off-set by the smallest occurring value in the dataset in order to avoid taking the log of zero, and z-scored. Methylation d(CpG) were b-values. The following linear models were used to compute the associations:

eQTL: lm(transcriptomics ∼ SNP + AGE + SEX + BMI + DIAB + genoPC1 + genoPC2 + genoPC3) meQTL: lm(CpG ∼ SNP + AGE + SEX + BMI + DIAB + genoPC1 + genoPC2 + genoPC3)

eQTM: lm(transcriptomics ∼ CpG + SNP + AGE + SEX + BMI + DIAB + genoPC1 + genoPC2 + genoPC3) A significance cut-off of p-value <5×10^−8^ was used.

### Genetic variation – omicsdata associations (omicsQTLs)

Omicsdata was inverse-normal scaled and residual were computed using the linear model “lm(Omicsdata ∼ sex + age + bmi + diab + genoPC1 + genoPC2 + genoPC3 + somaPC1 + somaPC2 + somaPC3)”. After QC, excluding non-autosomal SNPs, MAF<5%, HWE pvalue<10^−6^, or genotyping rate <98% (Suhre et al., 2017), 1,221,345 SNPs for 353 samples were available. Additive linear models using Plink version 1.9 (Chang et al., 2015) were computed. Genomic inflation was lambda<1.04 for all traits. All associations with p<5×10^−8^ were lumped, treating variants with R^2^ < 0.1 as independent(Suhre et al., 2017). Phenoscanner ((Kamat et al., 2019), accessed 9 April 2019) was used to annotate the sentinel variants with GWAS hits, metabolomics and proteomics QTLs, and genes encodes at the locus, using R^2^>0.8 (LD from EUR), and limiting associations to p-value < 5×10^−8^. Genetic variants were annotated to human genome build 37.

### Methylation – omicsdata association (omicsQTMs)

Residuals of methylation beta values (CpG) were computed using the linear model “lm (CpG ∼ AGE + SEX + BMI + DIAB + Gran + NK + CD4T + CD8T + Mono + Bcell + genoPC1 + genoPC2 + genoPC3)” and then z-scored. Saliva and urine metabolites were first normalized by saliva and urine osmolality, respectively. All omics variables were then inverse normal-scaled and residuals computed using the linear model “lm (Omicsdata ∼ AGE + SEX + BMI + DIAB + genoPC1 + genoPC2 + genoPC3)” and then z-scored. Association statistics were then computed using the linear model “lm(CpG_residual ∼ Omicsdata_residual)”. Associations reaching an ad hoc significance level of 5×10^−8^ were retained. CpG sites were annotated for gene names and CpG position relative to the genes using the Illumina provided HumanMethylation 450k annotation file.

### RNA expression – omicsdata association (omicsQTRs)

RNA expression data with less than 100 valid data points or median expression levels below 1 TPM were removed. Expression data was log-scaled, with all values off-set by the smallest occurring value in the dataset in order to avoid taking the log of zero, and z-scored. Saliva and urine metabolites were normalized by saliva osmolality and urine creatinine obtained from the respective platform, respectively. The omicsdata was then inverse-normal scaled. Metabolites and then samples with more than 50% missing values were removed. Association statistics were then computed using the linear model “lm(OMICS ∼ transcriptomics + AGE + SEX + BMI + DIAB + genoPC1 + genoPC2 + genoPC3 + CD8 + CD4 + NK + Bcell + Mono + Gran + Eos)”. Associations reaching an ad hoc significance level of 5×10^−8^ were retained.

### Disease/trait associations from the GWAS catalog

We downloaded the GWAS catalogue (gwas_catalog_v1.0.2-associations_e100_r2021-01-14.tsv) and identified 6,694 variants that are in LD (r2>0.8) with one of the 587 sentinel SNPs (incl. the SNPs themselves). We then identified 2,294 records in the GWAS catalogue that reported on one of the 6,694 SNPs. Where multiple associations with a same trait were reported for a same locus, we kept only the strongest association.

## Data Availability

All data produced are available online at:

https://github.com/karstensuhre/comics

http://comics.metabolomix.com

## Graphical Abstract

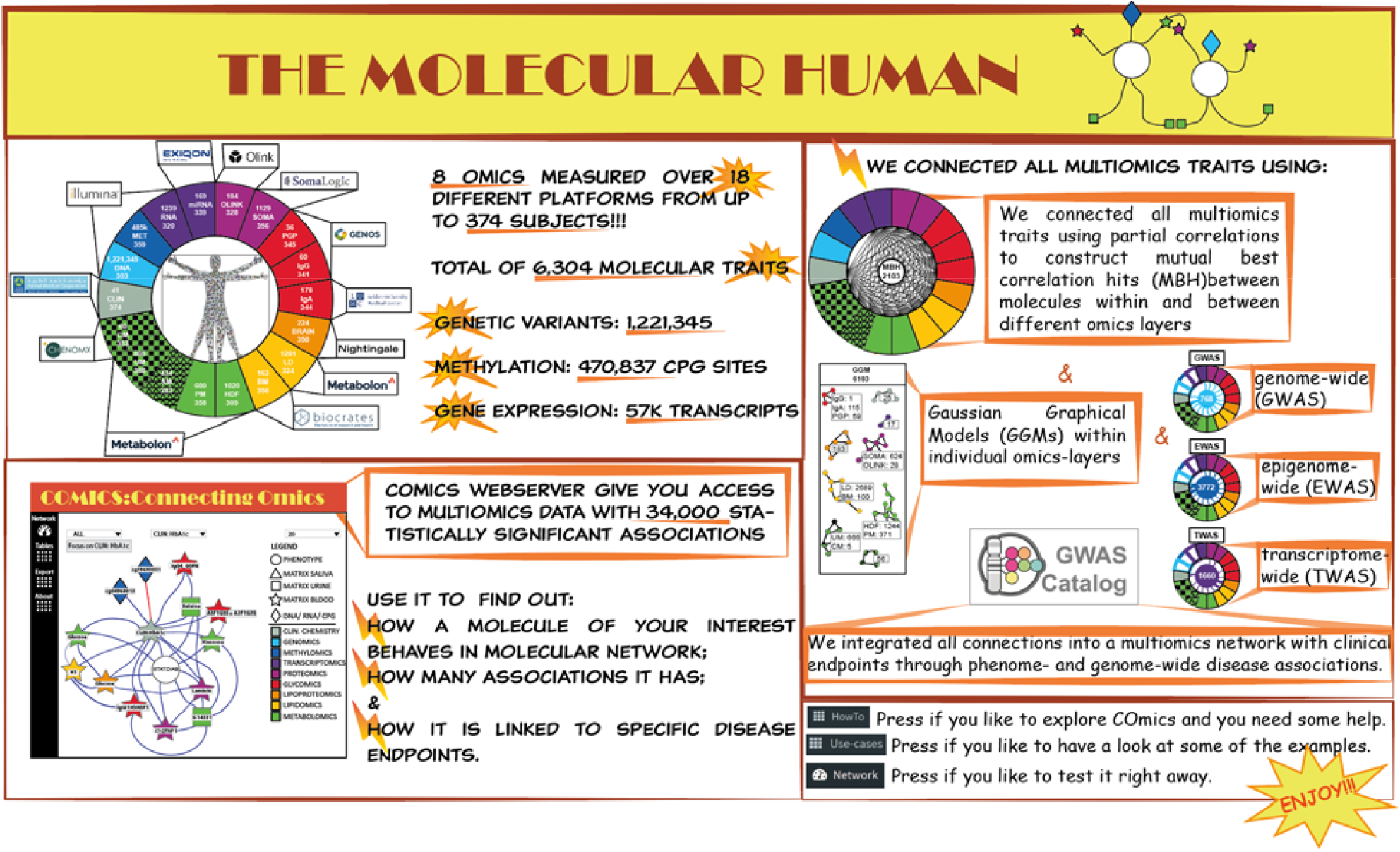

Understanding of omics complementarity, molecular interactions within and between various omics layers, and their link as interactive network into complex diseases is limited. With the current study we addressed this gap by deploying 18 technically diverse deep molecular phenotyping (omics-) platforms analyzing urine, blood, and saliva samples from up to 391 participants of the multi-ethnic diabetes case-control study QMDiab. We integrated quantitative readouts of 6,304 molecular traits with data on 1,221,345 genetic variants, DNA methylation at 470,837 CpG sites and gene expression of 57k transcripts into a molecular network including over 34,000 statistically significant associations. We constructed COmics (Connecting Omics) server (http://comics.metabolomix.com) to enable dynamic interaction with the data. With this study we draw Molecular human and COmics is a tool providing access into it.

## Text BOX

### BOX 1.

**Molecular networks related to T2D generated with COmics.**

We analyzed various T2D diabetes phenotypes using our COmics webserver and created a network around this phenotype (http://comics.metabolomix.com/?focus=STAT:%20DIAB). This network consists of 129 molecules (**Supplementary Table S5**) of which the majority were previously associated with T2D. Comics aids in the exploration of molecular interactions for individual molecules by the creation of a separate network centered on it.

We then analyzed the molecular network of HbA1C, which is considered a marker for long-term glycemic control: http://comics.metabolomix.com/?focus=CLIN%3A%20HbA1c%20(%25)&maxnodes=1 & we also investigated the adiponectin network a hormone possessing insulin-sensitizing properties and capacity for regulation of glucose and lipid metabolism: http://comics.metabolomix.com/?focus=SOMA%3A%20ADIPOQ%20%3A%20Adiponectin&maxnodes=1.

The adiponectin was previously found to be decreased in subject with T2D (Tabák et al., 2012). We identified multiple molecular associations across various omics levels with apparent relevance to the processes related to HbA1C and adiponectin as outlined in **Supplementary Information Note 9**. Interestingly, both networks revealed previously unreported associations. For instance, the glycans IgGI1H3N5F1 and IgG4-G0FN were linked with HbA1C, indicating a potential involvement in glucose metabolism. The association identified between adiponectin and phospholipid transfer protein (PLTP) [OLINK], previously identified as regulator of HDL metabolism (Huuskonen et al., 2000)and recognized as an emerging cardiometabolic risk factor in T2D patients (Dullaart et al., 2012), further suggest potential interaction between those two proteins in the regulation of lipid metabolism.

**These examples confirm the validity of the generated network, ensuring server functionality in describing molecular interactions related to T2D**.

### BOX2.

**Further insight into molecular network of tumor biomarker (rs17271883 near FUT6) revealed by COmics.**

The genetic rs17271883 near FUT6 was identified as a tumor biomarker (GWAS catalogue p-value = 5.0 ×10) (He et al., 2014). We used COmics to assembled the associated molecular network and explore the underlying mechanisms: http://comics.metabolomix.com/?focus=DNA%3A%20rs17271883%3Achr19%3A5834212%20FUT6&maxnodes=1 This network revealed direct genetic association between rs17271883 and two proteins detected on the SOMA platform including 3-galactosyl-N-acetylglucosaminide 4-alpha-L-fucosyltransferase (FUT3) (p-value = 1.2×10), and 4-galactosyl-N-acetylglucosaminide 3-alpha-L-fucosyltransferase (FUT5) (p-value = 3.8×10), as well as two CPGs including cg25387410 near FUT3 (p-value = 6.0×10) and cg16859884 (p-value = 3.2×10) in its close proximity. Fucosyltransferase (FUT) family members transfer fucose from GDP-fucose to sugar chains of glycoproteins or glycolipids (oligosaccharides). The family members FUT3, FUT5 and FUT6 share more than 85% homology on the genomic level and encode three similar fucosyltransferases (Reguigne-Arnould et al., 1995). This may explain the observed associations to rs17271883. Interestingly, expression of FUT3, FUT5 and FUT6 were all related to cancer occurrence and metastasis (Ej et al., 2015) and shown to be critical for cancer cell proliferation, migration and invasion (Liang et al., 2017). The Cpg’s site cg16859884, which we found associated with rs17271883, was also previously described in the context of cancer. Differences in cg16859884 methylation between malignant and normal prostate tissue was reported (Aref-Eshghi et al., 2018). Methylation site cg25387410 near FUT3 was not previously reported in the context of cancer, but given its placement in the molecular network presented here, its potential regulatory role in tumor pathogenesis may render it a promising candidate for further exploration. Increasing the number of nodes in the rs17271883 network revealed proteins involved in immune regulation including immunoglobulin lambda constant 2 (IGLC2), leukocyte immunoglobulin-like receptor B (LILRB) 5, complement factor H related 5 (CFHR5), Fc Gamma Receptor IIIb (FCGR3B), and tumor necrosis factor receptor superfamily member 17 (TNFRSF17) significantly associated with various glycans (**Supplementary Information Note 10**). Among those glycans we found multiple fucosylated molecules, which was expected given the function of fucosyltransferase. **The rs17271883 molecular network thus revealed an axis between the genetic variants, proteins and glycans with potential relevance to the pathogenesis of cancer and the role of immune regulation in the disease pathology more specifically**.

Another example pointing towards immune regulation based on the network constructed around rs103294 near AC010518.3 is described in **Supplementary Information Note 11**.

### BOX 3.

**Molecular network generated by COmics shed a light on the interplay between smoking, inflammation driven cardiovascular disorders and glycans.**

Platelet activation is enhanced in patients with myocardial infarction and is a hallmark of acute coronary syndrome (Frossard et al., 2004). Recent study established a direct link between smoking-induced hypomethylation of F2R like thrombin or trypsin receptor 3 (F2RL3) and platelet activity, providing further evidence for its contribution to smoking related cardiovascular risk (Corbin et al., 2022). Here, using the COmics webserver, we investigate the molecular network around the CpG cg05575921 at aryl hydrocarbon receptor repressor (AHRR) locus, a well-known smoking marker (Zaghlool et al., 2018), that is also significantly associated with arteriosclerosis, which is driven by inflammation, and carotid plaque (Ammous et al., 2022; Zhang et al., 2017). In depth investigation of this network http://comics.metabolomix.com/?focus=CPG%3A%20cg05575921%3Achr5%3A426378%20AHRR&maxnodes=1 is provided in **Supplementary Information Note 12**.

**The generated molecular network suggests an involvement of glycans in the link between smoking and inflammation driven cardiovascular disorders with G0FB glycan as a particularly auspicious candidate**.

## Supplementary Data

**Supplementary Table 1**. List of non-genomic molecular traits measured along with list of RNA deployed to conduct MBH

**Supplementary Table 2**. Molecules associated with age **Supplementary Table 3**. Molecules associated with sex **Supplementary Table 4**. Molecules associated with BMI **Supplementary Table 5**. Molecules associated with T2D

**Supplementary Table 6**. Correlation levels of statistically significant MBH.

**Supplementary Table 7**. Correlation levels of molecules measured on both SOMA and OLINK

**Supplementary Table 8**. GGM

**Supplementary Table 9**. Associations between gene SNPs and methylation levels (meQTL’s)

**Supplementary Table 10**. Associations between methylation levels and mRNA (eQTM’s)

**Supplementary Table 11**. Associations between gene SNPs and mRNA (eQTL’s)

**Supplementary Table 12**. Multiomics GWAS

**Supplementary Table 13**. Multiomics EWAS

**Supplementary Table 14**. Multiomics TWAS

## Acknowledgements

[please add as appropriate]

