## Supplemental Notes 1-12 for "The Molecular Human – A Roadmap of Molecular Interactions Linking Multiomics Networks with Disease Endpoints"

**Supplementary Information Content:**

**Notes:**

**Supplementary Note 1|** Proteins depicted as identical by SOMA and OLINK but showing low correlation differ in their molecular clusters.

**Supplementary Note 2|** Evaluation of platform performance through the strength of GWAS hits.

**Supplementary Note 3|** Current study replicate previous GWAS, EWAS and TWAS

**Supplementary Note 4|** Biological processes captured by MBH.

**Supplementary Note 5|** Linking SNP genotype, DNA methylation and gene expression.

**Supplementary Note 6|**GWAS additional findings.

**Supplementary Note 7|** EWAS additional findings.

**Supplementary Note 8|** TWAS additional finding.

**Supplementary Note 9|** Molecular network of T2D

**Supplementary Note 10|** Molecular network of rs17271883 related to cancer

**Supplementary Note 11|** Molecular network of rs103294 related to cancer

**Supplementary Note 12|** Molecular network related to cardiovascular disease

**Tables:**

**Supplementary Information Table 1|** The gene transcripts associated with CXCL11 and CXCL10 overlap with molecules involved in responses to viral infection.

**Figures:**

**Supplementary Information Figure 1|** Evaluation of proteins measured on both SOMA and OLINK which were not showing correlation and thy impact on molecular cluster

**Supplementary Information Figure 2|** Evaluation of platform performance through the strength of GWAS hits

**Supplementary Information Figure 3|**MBH between omics assembles molecular network related to biological processes.

**Supplementary Information Figure 4|** Example of association trios between the SNP-methylation, methylation-mRNA and SNP-mRNA.

**Supplementary Information Figure 5|** Example of EWAS associations with proteins measured on OLINK.

**Supplementary Information Figure 6|** Molecular network of cg05575921 methylation of aryl hydrocarbon receptor repressor (AHRR) and its implication in cardiovascular diseases

**Supplementary note 1.**

We investigated molecular networks around ICAM1 and CTSH measured on both SOMA and OLINK using COmics. We found different molecular networks around investigated molecules (**Supplementary Information Figure 1**). For instance, ICAM1 detected by SOMA was showing one GWAS association (rs923366 near ICAM1) and eight EWAS hits (e.g. cg03650189, cg04295144, cg08845333 near ICAM5) in contrast to OLINK which was showing association with alkaline phosphatase [CLIN], and phospholipids to total lipid ratio in medium HDL [BRAIN] (**Supplementary Information Figure 1A**). While investigating CTSH using COmics we found a cluster around CTSH measured by OLINK but not by SOMA (**Supplementary Information Figure 1B**). The OLINK network showed three GWAS (rs4778730 near CTSH, rs34593439 near CTSH, rs13345 near CTSH) and two EWAS (cg08500346 near CTSH, and cg17506061) hits. The identification of pQTL could be seen as validation of measurements specificity for targeted protein. Nevertheless, it should be noticed that in the aptamer/antibody techniques the binding affinity towards different isoforms might not reflect on the actual protein level as previously suggested (Raffield et al., 2020). For future experimental settings, the proteins showing low correlation should be validated with alternative technical platform.

**Supplementary Note 2**

We have investigated various GWAS associations to evaluate performance of different platforms. For instance, SNP rs1047891 associated with glycine with a p-value = 7.4 x10^-18^ in 320 samples on targeted lipidomics BM platform, with p-value = 7.4 x10^-15^ in 291 samples on the HDF platform, and with p-value = 7.1 x10^-14^ in 322 samples on the PM platform (**Supplementary Figure 2A**). In this example, the targeted assay appears to provide stronger signals, at least compared to the older PM platform, which was measured using an older generation of metabolic profiling described as HD2 by Metabolon. In another example however SNP rs1799958 associate with butyrylcarnitine (C4) with p-value = 7.9 x10^-9^ in 323 samples on targeted lipidomics BM platform, and p-value = 2.3 x10^-15^ in 294 on the HDF platform, and with p-value = 1.6 x10^-23^ in 325 samples on the PM platform, suggesting that platform performance might depend on the molecule properties and how well a given molecule is captured by each platform (**Supplementary Figure 2B**). We have observed similar trend across other platforms including measurements of urine metabolome with CM and UM (for SNP rs9922704 with 3-hydroxyisovalerate (**Supplementary Figure 2C**)) and with plasma proteome SOMA and OLINK (for rs3896287 with LILRB2 (**Supplementary Figure 2D**); for SNP rs8176693 with TIE1 (**Supplementary Figure 2E**); for SNP rs9892586 with CCL14 (**Supplementary Figure 2E**)).

**Supplementary Note 3**

We were able to replicate multiple hits including GWAS associations in blood with metabolites at the PYROXD2, NAT8, ACADS, NAT2, AGXT2, UGT1A4, CPS1, NAT16, and NAT16 loci (Gieger et al., 2008; Illig et al., 2010; Long et al., 2017; Shin et al., 2014; Suhre et al., 2011; Yu et al., 2014); with proteins including ACP1, ICAM1, FCGR2A, IL6R, ABO, SIGLEC9, ACP6, ENPP7, CCL15, PDGFRB (Di Narzo et al., 2017; Emilsson et al., 2018; Gilly et al., 2020; Suhre et al., 2017); with glycans including FUT6, ST6GAL1, MGAT5, C1GALT1 (Huffman et al., 2011; Kiryluk et al., 2017; Sharapov et al., 2019); and in the urine PYROXD2 and AGXT2 (Raffler et al., 2015; Schlosser et al., 2020); and in saliva one genetic loci SLC2A9 reported in the first and so far, the only one GWAS metabolomics study in saliva (Nag et al., 2020).

We also replicated multiple EWAS hits including cg07839457 near NLRC5, cg08122652 near PARP9 cg05575921 near AHRR, cg22910295 near ICAM5, cg13028630 near (*C4B*/*C4A*), and cg09488502 near SIGLEC4 (Bonder et al., 2017; Zaghlool et al., 2020).

Almost all of our TWAS with gene-traits replicated previous findings (**Supplementary Table 6**).

**Supplementary Note 4.**

The clinical chemistry parameters were showing largest number of MBH with metabolomics (45), proteomics (28) and glycomics (17). The significant associations identified between the CLIN and omics platforms were depicting identical molecules e.g. urate (HDF) ⬄ Uric acid (umol/L) (CLIN) (p-value = 6.71x10^-125^; r = 0.91); urate (PM) ⬄ uric acid (umol/L) (CLIN) (p-value = 6.71x10^-125^; r = 0.91), or molecules, which are known to interact in the same process. For example, HbA1C [CLIN], known marker for diagnosis and monitoring of Type 2 Diabetes (T2D) (Jeffcoate, 2004; Rahbar et al., 1969), was showing association with the elevated blood glucose level measured on different platforms as well as other molecules previously described in the context of diabetes including betaine (Lever et al., 2012), mannose (Mardinoglu et al., 2017) and X-14331 (Yousri et al., 2015) **Supplementary Information Figure 4 A.**

The largest number of MBH across two different omics was found between lipidomics and metabolomics (317), proteomics and metabolomics (153), glycomics and metabolomics (71), proteomics and lipidomics (70), glycoproteomics and metabolomics (68) as well as proteomics and glycomics (59). Some of those MBH’s reflect on previously described molecular process, and other could be considered as new associations. The MBHs detected between Apolipoprotein E (APOE), involved in lipid metabolism, and different lipid molecules across various platforms (**Supplementary Information Figure 4 B**) e.g. APOE (SOMA) ⬄ Total cholesterol in VLDL (BRAIN) (p-value = 1.9x10^-38^; r = 0.65); APOE (SOMA) ⬄ Total [FA16:0] (LD) (p-value = 4.2x10^-37^; r = 0.62); APOE (SOMA) ⬄ palmitoyl-linoleoyl-glycerol (16:0/18:2) (HDF) (p-value = 5.5x10^-20^; r = 0.58); APOE (SOMA) ⬄ 1-palmitoylglycerol (1-monopalmitin) (PM) (p-value = 1.9x10^-19^; r = 0.49); APOE (SOMA) ⬄ PC.aa.C34.2 (BM) (p-value = 1.0x10^-11^; r = 0.34), further suggest that actual biological processes can be captured by the MBH.

The majority of MBH identified between proteomics and glycomics replicated the associations reported by us previously (Suhre et al., 2019). The MBH’s between proteomics and transcriptomics showed frequently the gene transcripts and corresponding proteins SIGLEC14 (RNA) ⬄ SIGLEC14 (SOMA) (p-value = 1.1x10^-37^; r = 0.60); GNLY (RNA)⬄ GNLY (SOMA) (p-value = 9.6x10^-22^; r = 0.49); LILRA5 (RNA)⬄ LILRA5 (OLINK) (p-value = 2.0x10^-9^; r = 0.33).

Taken together, this data indicate that associations depicted by the MBH reflect on the actual biological processes and further pinpoint the complementarity between platforms describing two different omics.

**Supplementary Note 5**

We have identified total of 1,381 (meQTL’s) associations between gene SNPs and methylation levels, 15,991 (eQTM’s) associations between methylation levels and mRNA and 17 (eQTL’s) associations between gene SNPs and mRNA that reached a significance level of p<5x10^-8^. The meQTL’s consist of 203 unique SNPs and 308 unique methylation sites among which prolific SNPs (e.g. rs8283 near ATF6B associated with 33 methylation sites; rs4251552 near IRAK4 associated with 21 methylation sites, or rs12924274 near CNTNAP4 with 19 methylation sides) and prolific methylation sites (e.g. cg26831081 and cg05877118 near CLYBL with 75 and 73 SNPs, respectively; or cg03955321 near NDUFAF1 with 63 SNPs) were identified. Among eQTM’s we identified 280 unique methylation sites and 239 unique mRNA’s where we found many prolific methylation sites (e.g. cg04968013 near CACNB2 associated with 216 mRNA’s, cg08437802 near KANK2 associated with 205 mRNA’s) and mRNA’s (e.g. PRSS33 associates with 764 methylation sites, SIGLEC8 associates with 673 methylation sites or CLC associated with 665 methylation sites). The 17 identified eQTL’s included 12 unique SNP’s and 7 unique mRNA’s. We have identified two prolific mRNA’s including C4B and C4A associated with 6 and 5 SNP’s, respectively. We also identified association trios between the SNP-methylation, methylation-mRNA and SNP-mRNA (**Supplementary Information Figure 5**).

**Supplementary Note 6**

The multiomics GWAS conducted with all omics phenotypes measured across all platforms resulted in identification of 768 omicQTLs at 586 independent genetic loci that reached a significance level of p<5x10^-8^. We replicated majority of them and found 264 previously unreported in the PhenoScanner database of human genotype-phenotype associations (Kamat et al., 2019).

The metabolomics GWAS hits which we have identified as previously unreported included 42 plasma metabolites (22 HDF and 20 PM platforms) and 36 urine metabolomics (21 with CM and 15 with PM). Some of the urine GWAS identified on the CM platform were prolific; these include e.g. galactose with 7 hits. The identified here urine GWAS hits including QTL near ALMS1, ACADS, NAT2, and RNU6-675P were previously reported in the blood (Lotta et al., 2021; Schlosser et al., 2020; Shin et al., 2014) but not in the urine.

We found 96 previously unreported protein GWAS hits (including 76 pQTLs with SOMA and 20 pQTLs with OLINK). Out of the top 10 previously unreported significant associations 7 were proteins measured on OLINK panel, which could be explained by the relatively limited number of studies deploying this technology. The previously unreported pQTLs with the strongest association was between rs616114 (near MEP1B and GAREM1 genes) and level of MEP1B (p-value = 2.1x10^-45^; beta = -0.88). The MEP1B is meprin β plasma membrane associated protein previously suggested to be involved in diabetic nephropathy (Imperatore et al., 2001). The MEP1B deficiency was shown to be associated with higher mortality rates and more severe diabetic kidney injury in mice with STZ-induced T1D (Bylander et al., 2017), and was shown that MEP1B impacts complications of diabetes such as diabetic kidney injury by altering distinct metabolite profiles (Gooding et al., 2019).

Out of 38 identified lipidomics-QTLs we have only replicated one previously reported namely association between rs1799958 and plasma butyrylcarnitine (p-value = 7.9x10^-10^; beta = 0.50) (Long et al., 2017). Among the remaining 37 associations some including 18 between rs4493662 (near CTB-95D12.1/EEF1GP2) and various different triacylglycerols (TAG55 – TAG58) and 6 between rs79659787 (near RAB37) and different triacylglycerols (TAG48 – TAG52) were prolific.

**Supplementary Note 7**

We have identified 77, 22, and 7 significant oQTM’s with OLINK, LD, and miRNA, respectively. The most prolific trait oQTL were among OLINK associations including TYMP (27 associations), ENPP7 (9 associations), FCGR3B (6 associations), and FCRL1 (6 associations). We found that two of CPG’s associated with FCGR3B (cg26435281, and cg26561570) were on the gene locus of the associated protein (**Supplementary Information Figure 6A**) and out of nine CpG’s associated with ENPP7 protein, seven were on the gene locus of associated protein and two in very near proximity on the same chromosome. The ENPP7 measured on OLINK showed MBH with ENPP7 measured on SOMA (p-value = 1.33x10^-103^, r = 0.83), which was associated with the same CpG’s (**Supplementary Figure 6B**). Additionally, the ENPP7 protein associate with SNP rs3923265 near ENPP7 (p-value = 2.8x10^-24^, beta = 0.67), which was previously reported (Sun et al., 2018). The ENPP7 (Ectonucleotide pyrophosphatase/phosphodiesterase family member 7) is an alkaline sphingomyelinase that hydrolysis sphingomyelin (SM) to ceramide and its reduced activity was detected in colorectal cancer and in familial adenomatous polyposis, further suggesting a link between SM accumulation and colon tumorigenesis (Borza et al., 2022). Here we are suggesting potential role of methylation in regulation of ENPP7 protein expression.

**Supplementary Note 8**

Across the multiomics TWAS we have identified prolific associations between molecule (e.g. protein) and multiple gene transcripts. For instance, Mullerian-inhibiting factor (AMH) was associated with 195 different transcripts, P-selectin (SELP) with 18 transcripts, C-X-C Motif Chemokine Ligand 10 (CXCL10), and C-X-C Motif Chemokine Ligand 11 (CXCL11) with 16 and 6 transcripts, respectively. The CXCL10 and CXCL11 are inflammatory chemokines, induced by the interferon (IFN)-γ, regulating differentiation of naive T cells to T helper 1 (Th1) cells and migration of immune cells to their focal site (Tannenbaum et al., 1998). All the gene transcripts associated with CXCL11 overlap with the one identified as significantly associated with CXC10 and were previously reported as molecules involved in responses to viral infection (**Supplementary Information** **Table 1**).

**Supplementary Note 9**

The HbA1C molecular network consists of 14 molecules. Six out those 14 molecules were metabolites including glucose measured on multiple platforms, plasma mannose, urine betaine and urine X-14431. The glucose is a directly impacting HbA1C level (Nathan et al., 2007) and there is a clear interaction between glucose and mannose in physiological conditions as well as in pathology of T2D (Halama et al., 2020; Yoshimura et al., 2017). We identified two CpG’s namely cg19693031 (p-value = 4.9x10^-11^; TXNIP) and cg04968013 (p-value = 3.1x10^-8^; CACNB2) in the HbA1C molecular network. The TXNIP was shown as regulator of peripheral glucose metabolism (Parikh et al., 2007) and cg19693031 was previously associated with T2D and sustained hyperglycemia (Soriano-Tárraga et al., 2016). The CACNB2 was reported as susceptibility gene for diabetic retinopathy, which is a result of poor glycemic control (Wong et al., 2016), further suggesting potential role of cg04968013 in glucose metabolism. Among the molecules in HbA1C network, we also identified laminin, previously implicated in diabetes etiology and glucose metabolism (Antinozzi et al., 2006) and Complement C1q tumor necrosis factor-related protein 1 (C1QTNF1) determined as protective factor against diet-induced hyperglycemia accelerating fatty acid and glucose oxidation (Han et al., 2016). In the network we have also identified three glycans (IgGI1H3N5F1, IgG4-G0FN, and PGP29 (A3F1G3S(3,3,3)3 +A3F1G3S(3,3,6)3)); we previously reported on PGP29 in context of TXNIP association and T2D (Zaghlool et al., 2018) yet the identified association between PGP29 and HbA1C further suggest its role in T2D by involvement in glucose metabolism.

The molecular network of adiponectin shows negative associated with T2D phenotype, and positive association with different metabolites predominantly lipid structures including high-density lipoprotein cholesterol (HDL-C) [CLIN], previously described as positively associated with adiponectin (Magge et al., 2011), phospholipids in very large HDL [BRAIN], three glycerophospholipids ([LD], [BM], [HDF]), as well as 3-methylcrotonyl glycine [UM]. Noteworthy, the HDL-C as well as phospholipids in very large HDL levels were shown to be decreased in T2D patients (Gordon et al., 2013; Haase et al., 2015), which critical role of adiponectin.

**Supplementary Note 10**

Among the molecules which were not directly associated with rs17271883 but creating the network we have identified 2 other SNP’s near FUT6 namely rs8101385, and rs3760775 as well as 54 different glycans. The rs3760775, similarly to rs17271883 was previously described as tumor biomarker (He et al., 2014), and rs8101385, which we found to be associated with multiple glycans, was previously linked with IgG glycosylation (Shen et al., 2017). Interestingly, role of glycans in cancer progression was suggested and glycans were recognized as treatment targets (Dube and Bertozzi, 2005). Noteworthy, the immune system can be recruited to target cancer cells on the basis of their altered glycosylation (Dube and Bertozzi, 2005). In the molecular network we have identified multiple proteins involved in immune regulation including immunoglobulin lambda constant 2 (IGLC2), leukocyte immunoglobulin-like receptor B (LILRB) 5, complement factor H related 5 (CFHR5), Fc Gamma Receptor IIIb (FCGR3B), and tumor necrosis factor receptor superfamily member 17 (TNFRSF17) which were significantly associated with various glycans. The cancer relevance was previously reported for the proteins including IGLC2 (Bedognetti et al., 2015), LILRB5 (Carosella et al., 2021), FCGR3B (Treffers et al., 2019), and TNFRSF17 (Da Vià et al., 2021) as well as Leucine Rich Repeats And Immunoglobulin Like Domains 1 (LRIG1) (Billing et al., 2021) identified in rs17271883 near FUT6 molecular network.

**Supplementary Information Note 11**

Another cancer related example could be presented for molecular network of rs103294 near AC010518.3, which was identified as cancer-risk SNP (Fagny et al., 2020) associated with prostate cancer (GWAS catalogue p-value = 5x10^-16^). Overall the network around rs103294, with the maxim of 200 nodes, consist of 53 associations. The molecules associating directly with rs103294 includes methylation of DNA cg13762704 near LILRB1 [p-value = 5.5x10^-71^] as well as LILRB2 proteins measured on SOMA [p-value = 2.9x10^-18^] and OLINK [4.1x10^-10^]. The association between rs103294 and LILRB1 (cg13762704) methylation as well as LILRB2 protein was previously reported (Gaunt et al., 2016; Sun et al., 2018). The LILRB family possess immunosuppressive functions, and its family members were considered as immune checkpoint factor (Kuroki et al., 2019) with potential role in cancer cell immune evasion (Carosella et al., 2021; Chen et al., 2012).

**Supplementary Information Note 12**

We found direct association between cg05575921 AHRR methylation and 7 metabolites (**Figure 7B**) including cotinine N-oxide (p-value = 3.5x10^-13^), 3-methyl catechol sulphate 1 (p-value = 1.2x10^-9^), 3-methyl catechol sulphate 2 (p-value = 6.6x10^-10^), 2-ethylphenylsulfate (p-value = 7.7x10^-10^), 3-ethylphenylsulfate (p-value = 7.1x10^-14^), o-cresol sulfate (p-value = 2.2x10^-19^), and 2-naphthol sulfate (p-value = 2.3x10^-9^) which were previously described as markers of smoking (Gu et al., 2016). The cg05575921 AHRR methylation also showed association with protein detected on SOMA polymeric immunoglobulin receptor (PIGR) (p-value = 2.2x10^-9^), previously identified as significantly elevated in plasma of smokers and associated with smoking cigarettes (Ohlmeier et al., 2012). The cg05575921 AHRR methylation showed association with G-protein coupled receptor 15 (GPR15) (p-value = 2.2x10^-9^). Interestingly, previous study reported increased expression of GPR15 in smokers especially in the T-cells of smokers and inflammation (Andersen et al., 2021), further suggesting its previously unrevealed potential role in pathology of atherosclerosis. The significant portion of the network constitute of IgA glycans, and there are nine glycans directly associated with cg05575921 AHRR methylation. The modifications in IgG glycan structures were previously attributed to smoking (Knežević et al., 2010) and AHRR methylation was previously reported as strongly associated with IgG glycans (Wahl et al., 2018) . Interestingly, out of the glycans associated with AHRR methylation, one G0FB (A2B) also associated with other CPG’s including two PARP9 methylations (cg00959259 and cg22930808), CMPK methylation (cg01028142), F2RL3 methylation (cg03636183), IFIT3 (cg06188083) methylation, IFI44L methylation (cg13304609), LY6E methylation (cg14392283) and IRF7 methylation (cg22016995) (Supplementary Information **Figure 7C**). Three out of those methylations (PARP, F2RL3 and IFIT3) were previously associated with smoking (Breitling et al., 2011; Sikdar et al., 2019), and seven (PARP, CMPK, IFIT3, IFI44L, LY6E and IRF7) with systemic lupus (Breitbach et al., 2020; Imgenberg-Kreuz et al., 2018; Joseph et al., 2019), which is a chronic autoimmune disease with cardiovascular comorbidities (Fava and Petri, 2019).

**Supplementary Information Table 1.** The gene transcripts associated with CXCL11 and CXCL10 overlap with molecules involved in responses to viral infection.

| RNA TRANSCRIPT ID | TRANSCRIPT NAME | DESCRIPTION | REF. |
| --- | --- | --- | --- |
| ENSG00000149131 | SERPING1 | SERPING1 was identified as important component of the innate immune system to restrict HIV-1 infection. Positive association between IFITM1-an antiviral molecule and SERPING1 was reported. | (Sanfilippo et al., 2017) |
| ENSG00000119917 | IFIT3 | IFIT3 is an IFITs family member involved in immune responses and restrict viral infections through a variety of mechanisms, including the restriction of viral RNA translation. IFIT3 modulates IFIT1 RNA binding specificity and protein stabilit. | (Johnson et al., 2018) |
| ENSG00000134321 | RSAD2 | Rsad2, also known as viperin, is an interferon-stimulated gene of innate immunity involved in antiviral responses implicated in inhibition of broad spectrum of DNA and RNA viruses. | (Chin and Cresswell, 2001; Gizzi et al., 2018) |
| ENSG00000133106 | EPSTI1 | Epsti1 is highly expressed in macrophages exposed to IFNγ and lipopolysaccharide (LPS) and was recognized as a modulator of macrophage activation and polarization. | (Kim et al., 2018) |
| ENSG00000168062 | BATF2 | Batf2 is induced IFN-γ-activated classical macrophages and was reported to play a role in innate immune response by controlling expression of several important immune regulatory genes, such as Nos2, Tnf, Ccl5, Cxcl9, Cxcl11, Ccr5, Cxcr3, Il6, and Niarc1, by forming the Batf2/Irf1 complex | (Roy et al., 2015) |
| ENSG00000134326 | CMPK2 | Mitochondrial CMPK2 mediates immunomodulatory and antiviral activities through IFN-dependent and IFN-independent pathway and is known to interplay with RSAD2. | (Lai et al., 2021) |
| ENSG00000117228 | GBP1 | GBP1 is a member of Guanylate-binding proteins (GBPs) proteins family inducible by the IFNγ. The GBP1 was reported as microbe-specific gatekeeper of macrophage apoptosis and pyroptosis. | (Fisch et al., 2019) |
| ENSG00000156587 | UBE2L6 | Ube2L6 is induced on the transcriptonal level by IFNγ and is a critical enzyme in ISGylation that conjugates the ubiquitin-like modifier, interferon-stimulated gene 15 (ISG15), to target substrates. | (Orfali et al., 2020) |
| ENSG00000187608 | ISG15 | ISG15 is a member of the ubiquitin family strongly induced by type I interferons which shapes the host response to viral infection. | (Perng and Lenschow, 2018) |
| ENSG00000185745 | IFIT1 | IFIT1 is a member of antiviral, RNA-binding protein family, which are among the highest expressed genes during antiviral immune response. IFIT1 recognizes 5′-triphosphate RNA activated under the viral infection. IFIT1 is a part of the the IFIT complex which antagonizes viruses by sequestering specific viral nucleic acids. | (Pichlmair et al., 2011) |
| ENSG00000126709 | IFI6 | Is a member of IFI, well-known family of cytokines with antiviral effects stimulating immune response by IFN-stimulated genes (ISG). IFI6 was reported as an ER-resident interferon effector that blocks flavivirus replication. | (Richardson et al., 2018) |
| ENSG00000137959 | IFI44L | IFI44L, an ISG induced by many different viruses, was defined as feedback regulator of host antiviral responses. It was shown that IFI44L gene is negatively modulating innate immune responses induced after virus infections and that decreasing IFI44L expression impairs virus production. | (DeDiego et al., 2019) |
| ENSG00000111331 | OAS3 | OAS3 is a member of 2'-5'-oligoadenylate synthetases (OAS) known to enhance intracellular antiviral mechanisms by stimulation of cytokine secretion. | (Leisching et al., 2019) |
| ENSG00000119922 | IFIT2 | IFIT2 is a member of antiviral, RNA-binding proteins family, and was reported as effector protein of Type I IFN–mediated amplification lipopolysaccharide (LPS)-induced cytokine production. | (Siegfried et al., 2013) |
| ENSG00000138646 | HERC5 | HERC5 is an IFN-induced HECT-type E3 protein ligase that mediates type I IFN-induced ISGylation of protein targets. | (Wong et al., 2006) |
| ENSG00000137965 | IFI44 | IFI44 expression is inducible by IFN-α and -β but not by IFN-γ. IFI44 was shown to suppresses HIV-1 LTR promoter further affecting viral transcription. | (Power et al., 2015) |

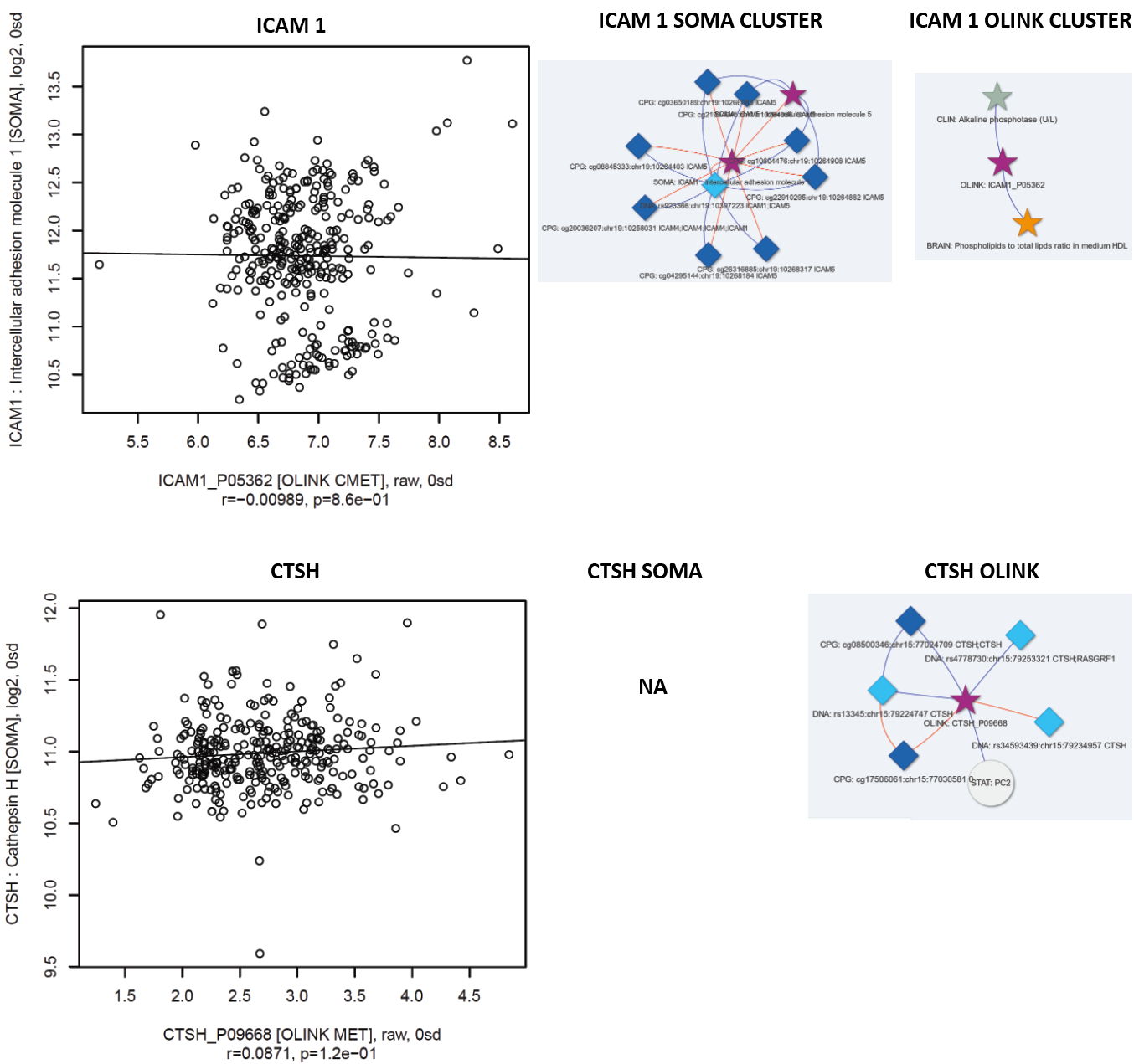

**Supplementary Information Figure 1.** Evaluation of proteins measured on both SOMA and OLINK which were not showing correlation and thy impact on molecular cluster. **A)** ICAM1 and **B)** CTSH.

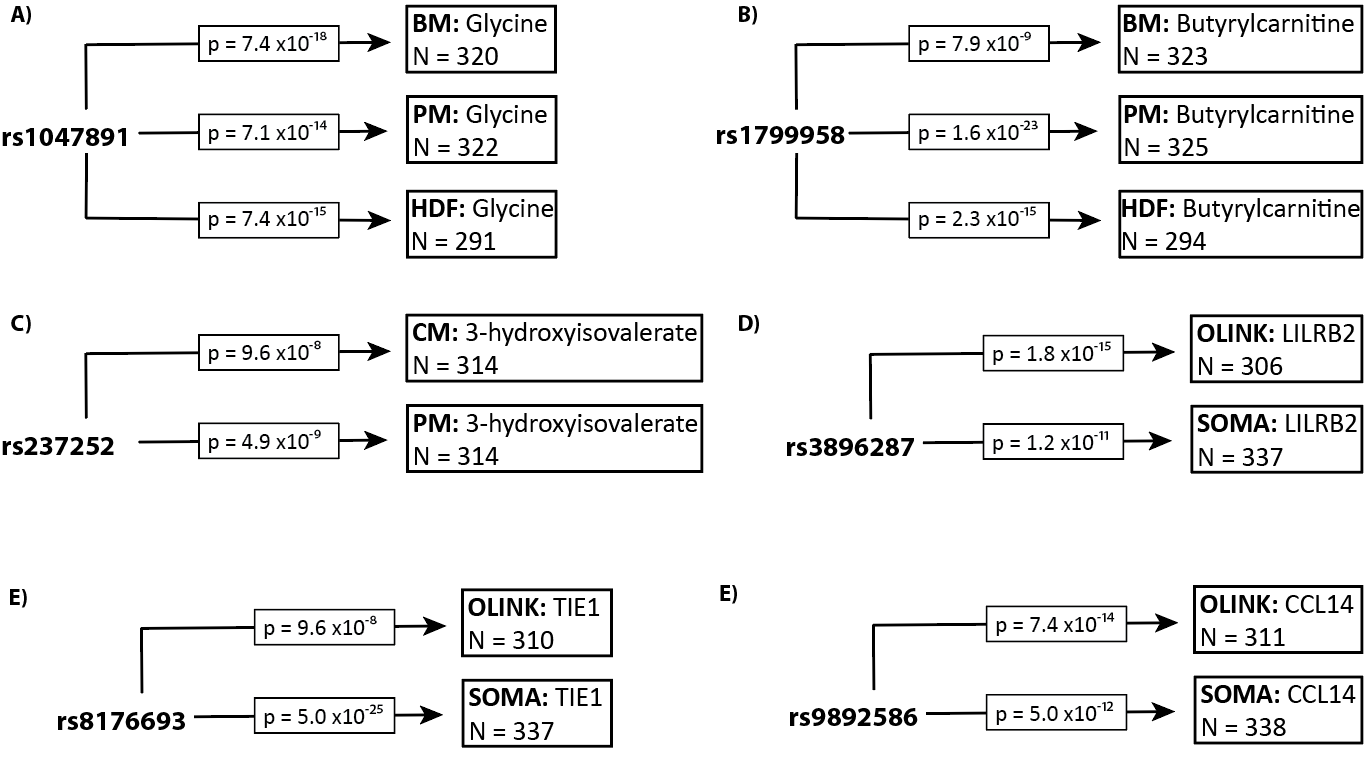

**Supplementary Figure 2. Evaluation of platform performance through the strength of GWAS hits. A)-E)** Multiomics GWAS hits.

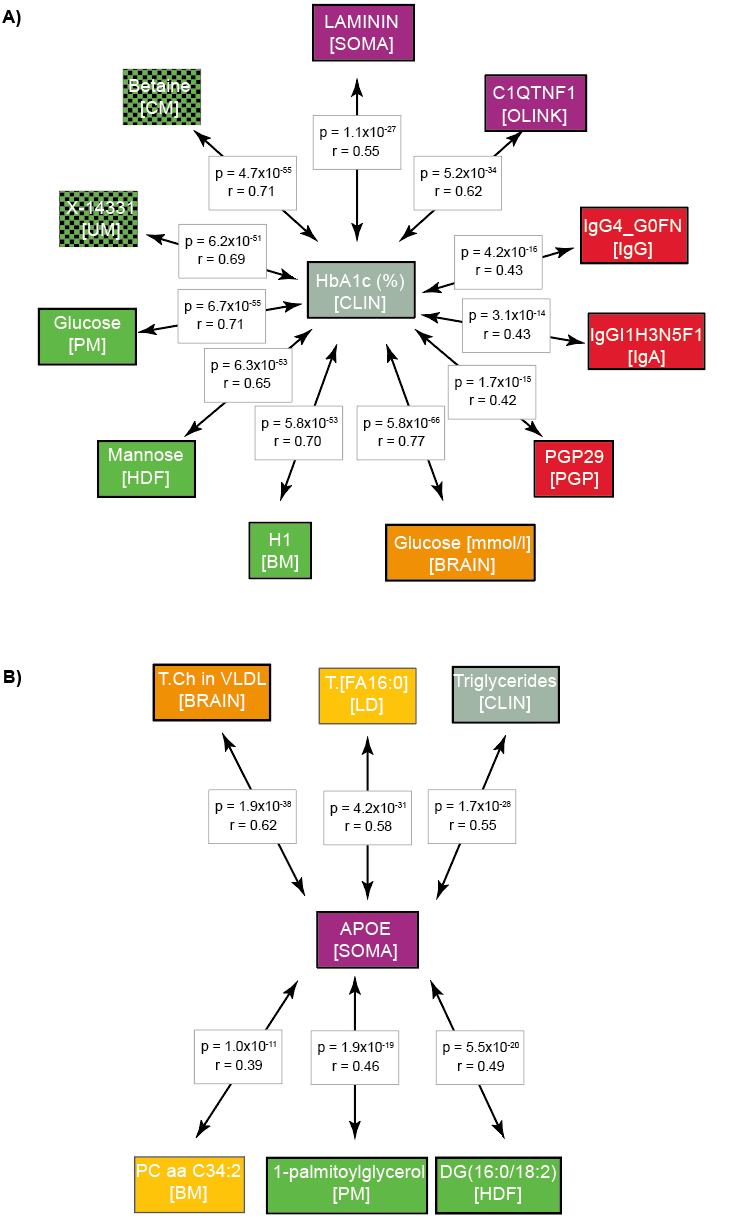

**Supplementary Information Figure 3**. **MBH between omics assembles molecular network related to biological processes. A)** MBH identifies clinically relevant association between HbA1C and molecules involved in pathology of T2D. **B)** MBH reflect on lipid metabolism.

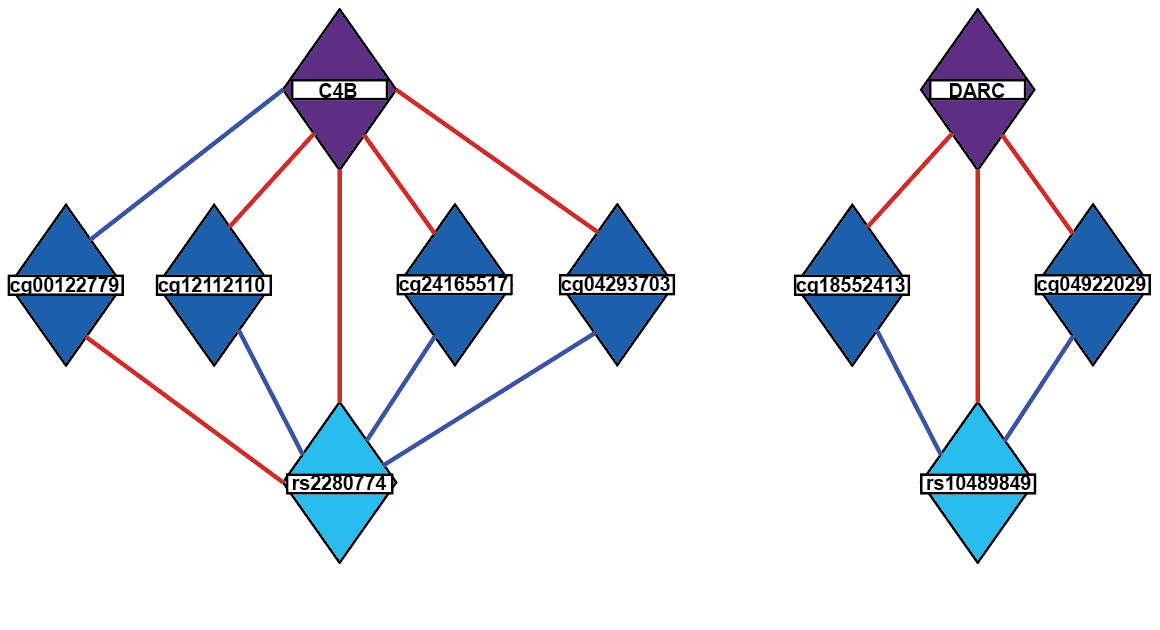

**Supplementary Information Figure 4.** Example of association trios between the SNP-methylation, methylation-mRNA and SNP-mRNA.

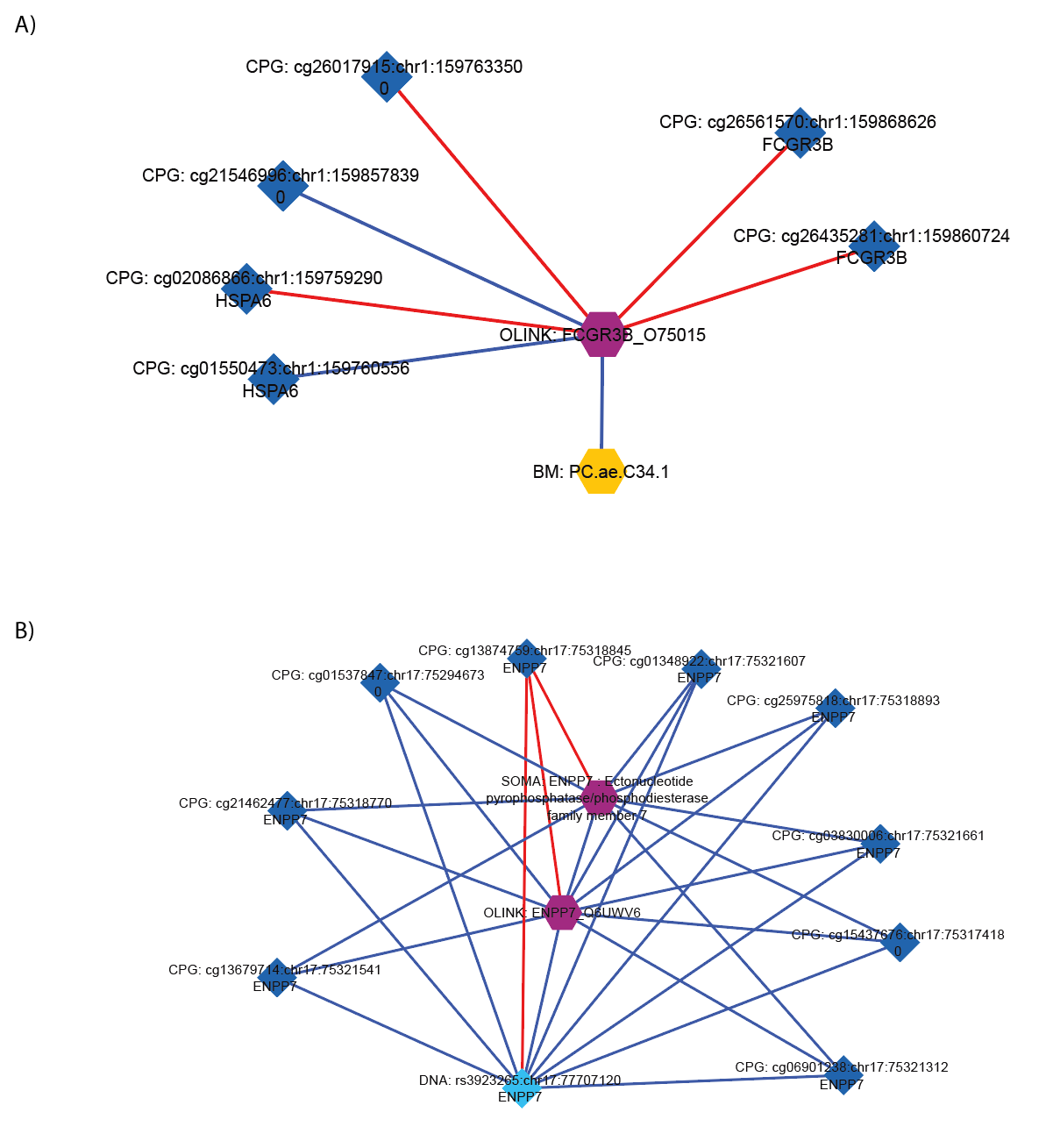

**Supplementary Information Figure 5**. Example of EWAS associations with proteins measured on OLINK.

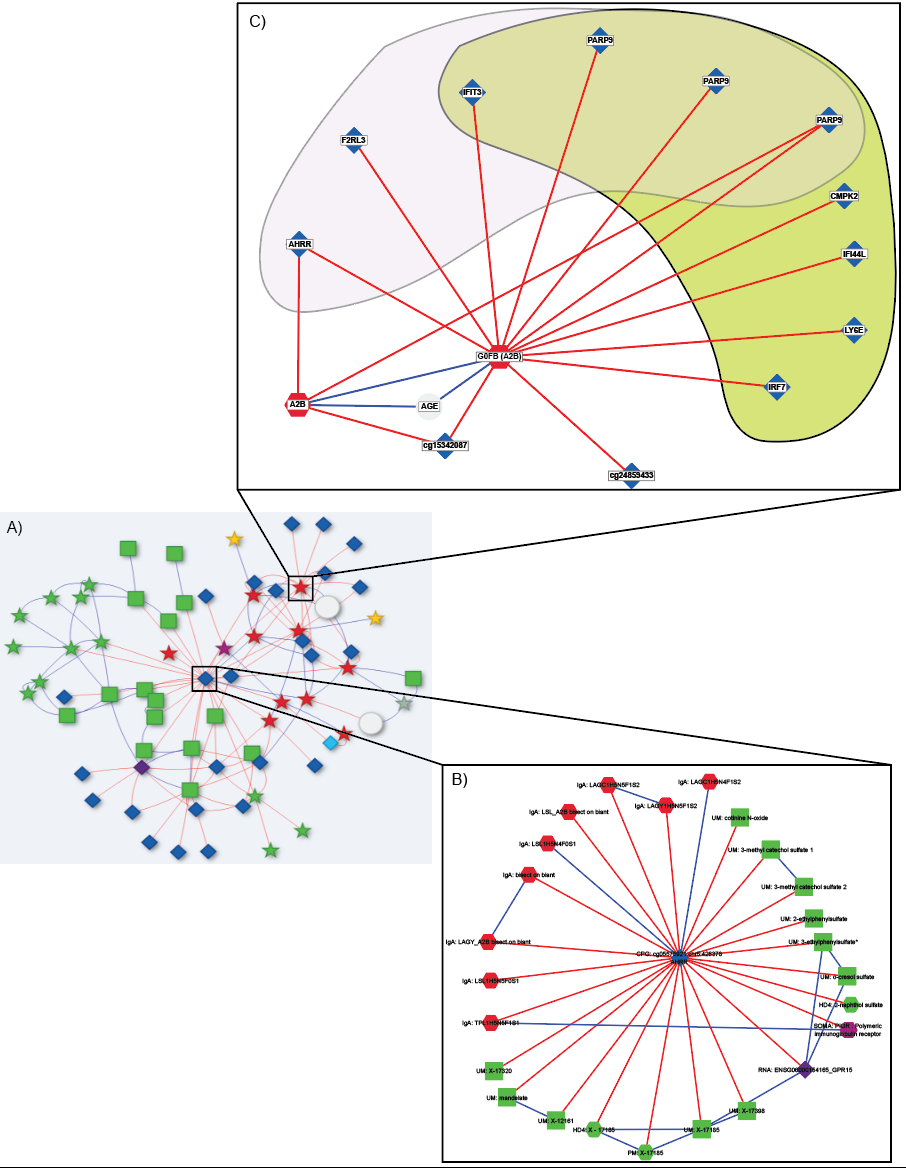

**Supplementary Information Figure 6. Molecular network of cg05575921 methylation of aryl hydrocarbon receptor repressor (AHRR) and its implication in cardiovascular diseases. A)** Global network of cg05575921. **B)** Molecules directly associated with cg05575921. **C)** CPG’s associated with G0FB (A2B) are regulated in response to smoking and systemic lupus.
